# Allergen recognition by specific effector Th2 cells enables IL-2-dependent activation of regulatory T cell responses in humans

**DOI:** 10.1101/2022.05.17.22275017

**Authors:** Daniel Lozano-Ojalvo, Scott R. Tyler, Carlos J. Aranda, Julie Wang, Scott Sicherer, Hugh A. Sampson, Robert A. Wood, A. Wesley Burks, Stacie M. Jones, Donald Y. M. Leung, Maria Curotto de Lafaille, M. Cecilia Berin

**Affiliations:** Jaffe Food Allergy Institute, Icahn School of Medicine at Mount Sinai; New York, NY; Precision Immunology Institute, Icahn School of Medicine at Mount Sinai; New York, NY; Genetics and Genomic Sciences, Icahn School of Medicine at Mount Sinai; New York, NY; Department of Pediatrics, Johns Hopkins University School of Medicine; Baltimore, MD; Department of Medicine and Pediatrics, University of North Carolina School of Medicine; Chapel Hill, NC; Department of Pediatrics, University of Arkansas for Medical Sciences and Arkansas Children’s Hospital; Little Rock, AR; Department of Pediatrics, National Jewish Health; Denver, CO

## Abstract

Type 2 allergen-specific T cells are essential for the induction and maintenance of allergies to foods, and Tregs specific for these allergens are assumed to be involved in their resolution. However, it has not been convincingly demonstrated whether allergen-specific Treg responses are responsible for the generation of oral tolerance in humans. We observed that sustained food allergen exposure in the form of oral immunotherapy resulted in increased frequency of Tregs only in individuals with lasting clinical tolerance. We sought to identify regulatory components of the CD4^+^ T cell response to food allergens by studying their functional activation over time *in vitro* and *in vivo*. Two subsets of Tregs expressing CD137 or CD25/OX40 were identified with a delayed kinetics of activation compared to clonally enriched pathogenic effector Th2 cells. Treg activation was dependent on IL-2 derived from effector T cells. *In vivo* exposure to peanut in the form of an oral food challenge of allergic subjects induced a delayed and persistent activation of Tregs after initiation of the allergen-specific Th2 response. Our results reveal a dependency of Tregs on effector Th2 cells for their activation and highlight the important role of IL-2 in the generation of a regulatory response to food allergens.

**One Sentence Summary:** Initiation of allergen-specific Th2 cell responses induces an IL-2-mediated activation of Tregs with suppressive properties.

## INTRODUCTION

Food allergies are thought to be the result of a defective regulatory T cell (Treg) function leading to the generation of food allergen-specific type 2 cellular immunity and IgE production ^1,2^. Current research efforts are focused on determining the clinical relevance of distinct allergen-specific T cell subsets not only in the generation and maintenance of this disease but also in its resolution. In humans, CD4^+^ T cell subsets contributing to maintenance of food allergies include allergen-specific Th2A, pathogenic effector Th2 (peTh2), and T follicular helper 13 (Tfh13) cells ^3-6^. These have been detected in peripheral blood and all are characterized by a poly-Th2 cytokine producing profile. Th2A and peTh2 cells express tissue-homing receptors while Tfh13 cells express CXCR5 for follicle homing. In addition, the frequency of circulating food allergen-specific Th2 cells has been related to important clinical phenotypes such as the threshold of reactivity to peanut allergens ^7,8^. Allergen-specific Th2 cells are reduced in response to oral immunotherapy (OIT), but it is unclear whether this treatment-induced reduction is a mechanism of food allergy resolution ^5,9,10^.

Although there is consistent evidence for the role of Tregs in the generation of oral tolerance to food allergens in mice ^11-13^, evidence in humans is lacking and the existence of circulating food allergen-specific Tregs remains controversial. In part, this may be due to the technology chosen to identify allergen-specific T cells. Peptide-MHC II tetramers may select for higher affinity MHC II-T cell receptor (TCR) interactions than those featuring Tregs ^14^. In allergies to airborne antigens, the activation markers CD154 (CD40L) and CD137 (4-1BB) are differentially expressed on allergen-specific effector CD4^+^ T cells (CD154^+^) and Tregs (CD137^+^CD154^-^) ^15,16^. However, this approach has not identified differences in peanut-specific Treg compartments between allergic and non-allergic subjects ^17^.

In this study, we hypothesized that in addition to the allergen-specific effector T cell responses induced by food allergens, regulatory T cells that oppose the Th2 pro-inflammatory effects are also generated. We observed that the expansion of Treg response during egg OIT was associated with the development of clinical tolerance after treatment with oral immunotherapy for egg allergy. To identify and functionally characterize previously unrecognized Treg responses, we chose to use an activation-based approach using a wide range of activation markers analyzed over time. We have determined that the early activation of allergen-specific peTh2 cells induced a cytokine-dependent activation of Tregs via IL-2 not only upon *in vitro* recognition of the peanut allergens but also after oral food challenge in peanut allergic subjects.

## RESULTS

### Egg OIT results in an expansion of Tregs associated with the induction of sustained oral tolerance in allergic subjects

We evaluated the frequency of CD127_low_CD25^+^ Tregs throughout the course of egg OIT in egg allergic individuals ^9,18^. Our results revealed a significant expansion of Tregs during OIT (Fig. 1A). Furthermore, when we analyzed these data based on the outcome of treatment, this augmented frequency of Tregs was only observed in the participants who reached sustained unresponsiveness (SU; Fig. 1B). Discontinuation of the treatment (for 10-12 weeks, from month 24 to month 26) significantly reduced the total number of Tregs (Fig. 1B), indicating that sustained allergen exposure was required to maintain the elevated Treg population. CD154 was the sole activation marker available for analysis in this cohort, which was not informative in identifying the allergen specificity of the increased Tregs. We hypothesized that other activation markers could reveal the recognition of allergen-specific Tregs.

**Fig. 1.**
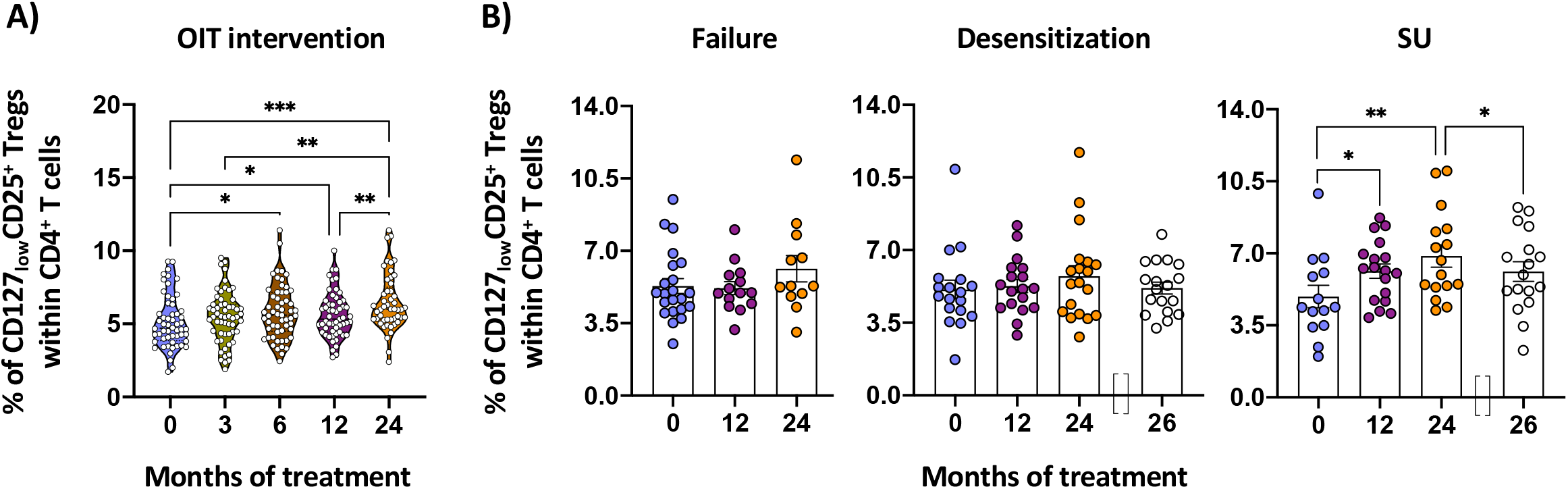
Oral immunotherapy (OIT) generates the expansion of regulatory T cells (Tregs) in those subjects who achieve sustained unresponsiveness (SU). **A**. Percentage of total CD127_low_CD25^+^ Tregs identified in PBMCs from egg allergic subjects following egg OIT (n=63). **B**. Induction of total Tregs throughout the course of egg OIT categorized by treatment resolution in: failure (left, n=24), desensitization (middle, n=21), and SU (right, n=18). Timepoints are baseline (T0), during treatment (3, 6, 12, and 24m), and 2m after discontinuation of the OIT (26m). Each data point is one individual in (A) and one individual (mean ± SEM) in (B). Statistical analysis by mixed-effect analysis with Geisser-Greenhouse correction followed by Tukey’s (in A) and Dunnett’s (in B) multiple comparisons tests. *P<0.05, **P<0.01, and ***P<0.001.

### Kinetics of memory CD4^+^ T cell activation in peanut allergic subjects reveals a delayed activation of allergen-induced Tregs

There is lack of information on how allergen-specific T cell responses evolve over time. We screened a set of markers previously described for tracking the early and late activation of effector and regulatory CD4^+^ T cells ^19^ by stimulating PBMCs from peanut allergic (PA) individuals with a crude peanut extract (CPE) and analyzing memory CD4^+^ T cells by spectral flow cytometry at three different timepoints (6, 24, and 48h). Unsupervised hierarchical clustering of the memory CD4^+^ T cells using the FlowSOM algorithm identified three meta-clusters based on expression of the activation markers CD154, CD137, OX40, CD69, and CD25 (Fig. 2A). These meta-clusters were characterized by enriched expression of CD137 in absence of CD154 (meta-cluster 1), CD154 and CD69 (meta-cluster 2), and CD25 and OX40 (meta-cluster 3) (Fig. 2B). We validated these three populations by manual gating (Fig. S1A). Peanut-induced CD154^+^CD69^+^ memory CD4^+^ T cells were increased early (6h) after stimulation compared to the unstimulated control (CTRL), and continued to increase after 24 and 48h (p<0.0001). By contrast, CD137^+^CD154^-^ and CD25^+^OX40^+^ T cells were significantly induced after 24h and their frequency was sustained at 48h (p<0.0001), thus showing a delayed activation compared to CD154^+^CD69^+^ T cells (Fig. 2C). To assess the specificity of these T cell responses, we examined two negative control groups that did not have peanut allergies: non-allergic and egg allergic individuals (Fig. S1B and S1C). None of these controls had significant peanut-induced T cell responses. In contrast, CD154^+^CD69^+^, CD137^+^CD154^-^, and CD25^+^OX40^+^ T cells were detected after 24h-stimulation of PBMCs from egg allergic subjects with egg white proteins (Fig. S1C).

**Fig. 2.**
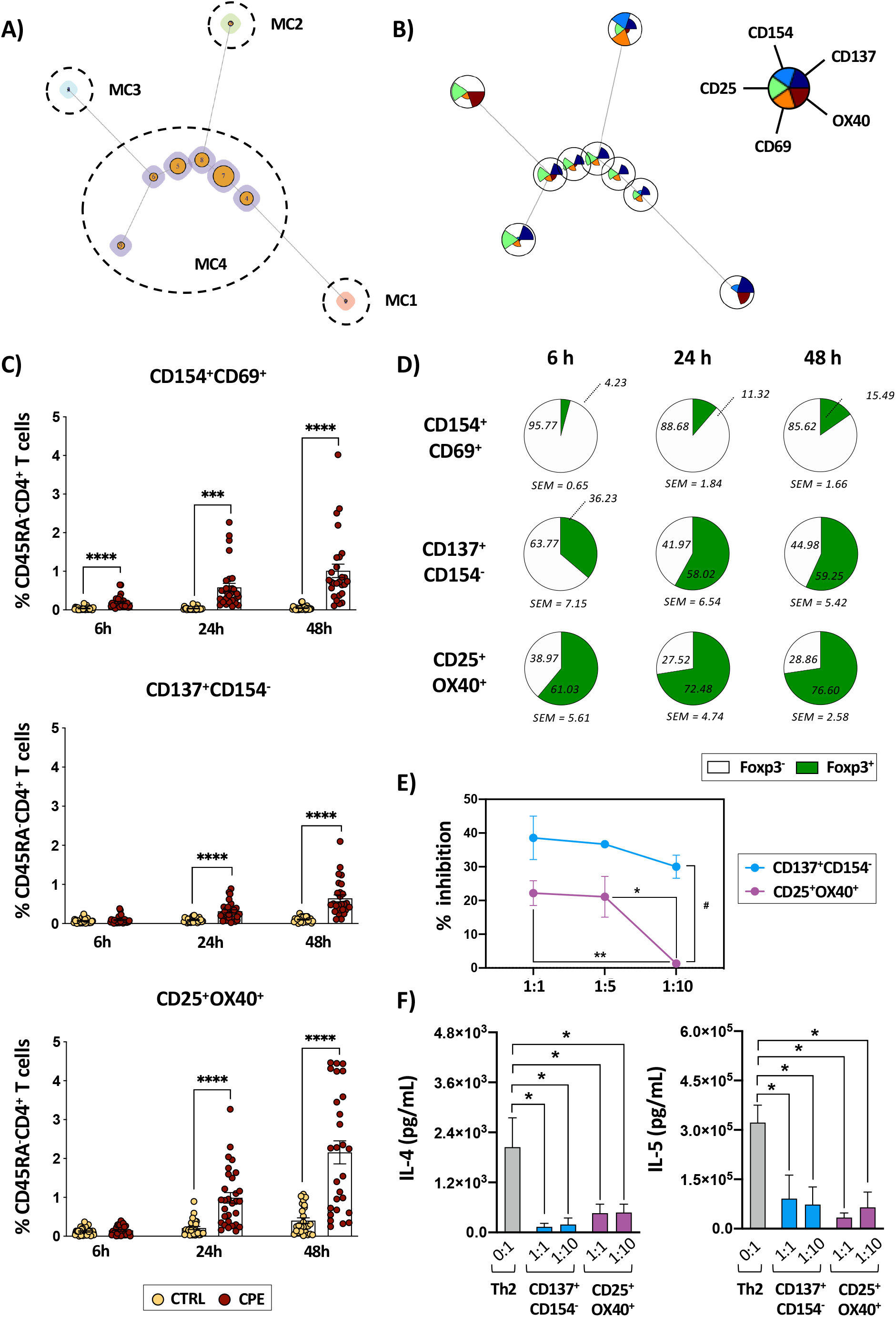
Peanut allergens induce an early T cell response driven by CD154^+^CD69^+^ cells and a delayed activation of suppressive regulatory T cells (Tregs) in peanut allergic (PA) subjects. **A**. Minimal spanning tree visualization of FlowSOM clustering analysis of memory (CD45RA) ^-^CD4^+^ T cells of PA subjects stimulated with crude peanut extract (CPE) for 6h, 24h, and 48h (n=5). Different nodes indicate the relative size of the cluster identified. Meta-clusters (MC) are indicated with different numbers. **B**. Intensity of expression of activation markers visualized by a star chart in each node of the FlowSOM clustering analysis (n=5). Each pie height indicates intensity of expression. **C**. Percentage of the activated populations identified in memory CD4^+^ T cells unstimulated (CTRL) or stimulated with CPE for 6h, 24h, and 48h (n=30). **D**. Percentage of Foxp3^-^ (white) and Foxp3^+^ (green) CD4^+^ T cells in the activated populations after CPE-stimulation for 6h, 24h, and 48h (n=13-14). **E**. Inhibition percentage of CD25^-^CD4^+^ T cell proliferation induced by activated Tregs populations sorted from polyclonally activated PBMCs. Suppression of CD25^-^ CD4^+^ T cell proliferation was calculated via CFSE dilution. **F**. Quantification of IL-4 and IL-5 released by polarized Th2 cells alone or cultured with activated Treg populations sorted from polyclonally activated PBMCs for 72h. In (C) each data point is one individual (mean ± SEM), each point represents the mean ± SEM of three independent experiments performed in triplicate in (E), and bar graphs represent mean + SEM of three independent experiments performed in triplicate in (F). Statistical analyses by mixed-effect analysis with Geisser-Greenhouse correction followed by Tukey’s multiple comparisons test in (C) and (E), and ordinary one-way ANOVA with Geisser-Greenhouse correction followed by Dunnett’s multiple comparisons test in (F). *P < 0.05, **P < 0.01, ***P < 0.001, and ****P < 0.0001.

To determine the presence of Tregs in these peanut-induced CD4^+^ T cell populations, we determined the Foxp3 expression over time (Fig. 2D). CD154^+^CD69^+^ cells were mainly Foxp3^-^ T cells at 6h after stimulation. In the peanut-induced CD137^+^CD154^-^ population we found an even distribution of Foxp3^-^ and Foxp3^+^ CD4^+^ T cells after 24h of CPE stimulation. By contrast, most of the activated cells contained in the CD25^+^OX40^+^ T cells expressed Foxp3 at all the timepoints studied (Fig. 2C; Fig. S2A). To better understand whether these populations of activated Tregs could be associated with a suppressive function *in vitro*, we generated these subsets by polyclonal T cell activation (Fig. S2C) and performed an immunosuppression assay using CD4^+^CD25^-^ responder T cells (Tresps) at different ratios (Treg:Tresp). Our results showed that CD137^+^CD154^-^ cells significantly suppressed Tresps proliferation already at a 1:10 ratio. CD25^+^OX40^+^ T cells, although containing a higher proportion of Foxp3^+^ cells (Fig. 2D), started exerting a suppressive activity at a ratio of 1:5 suggesting a more limited suppressive function of this population (Fig. 2E). To study their ability to suppress Th2 responses, we co-cultured polyclonally generated CD137^+^CD154^-^ and CD25^+^OX40^+^ T cells with Th2-polarized cells (Fig. 2F). Both activated populations of Tregs significantly reduced the levels of IL-4 and IL-5 secreted by Th2 cells at the ratios studied (Fig. 2F). Taken together, we have identified distinct kinetics of activation of two peanut-responsive Treg populations in PA subjects with suppressive activity on Th2 cells.

### Peanut-induced CD4^+^ T cell responses are characterized by an early activation of mature effector memory allergen-specific Th2 cells

Since activated CD4^+^ T cells can express several activation markers after TCR engagement in a time-dependent manner, we studied the overlap between the identified activated populations. Minor overlaps between populations were observed at all timepoints studied (Fig. 3A). We examined the clonotype diversity of the different peanut-induced T cell populations by sequencing the TCRβ chains and analyzing their complementarity-determining region 3 (CDR3). Our results revealed that early activated CD154^+^CD69^+^ T cells (6h) had a lower CDR3 sequence diversity and were highly enriched for specific clonotypes compared to the other populations analyzed (Fig. 3B; Fig. S3B and S3C). Tracking the CDR3 clonotypes that appeared in the early CD154^+^CD69^+^ at 6h within the other populations studied, we found a notable overlap within CD154^+^CD69^+^ T cells after 6h and 24h of CPE stimulation (Fig. S3C). In contrast, CD137^+^CD154^-^ and CD25^+^OX40^+^ T cells showed high CDR3 diversity consistent with less clonality (Fig. 3B; Fig. S3B and S3C) and a very little overlap between CD154^+^CD69^+^ 6h clonotypes and those found in these activated populations (Fig. S3C). We have identified distinct kinetics of activation of these three unique peanut-induced CD4^+^ T cell populations: early induced, clonally enriched CD154^+^CD69^+^ T cells (6h) and a delayed and less clonal response represented by CD137^+^CD154^-^ and CD25^+^OX40^+^ T cells.

**Fig. 3.**
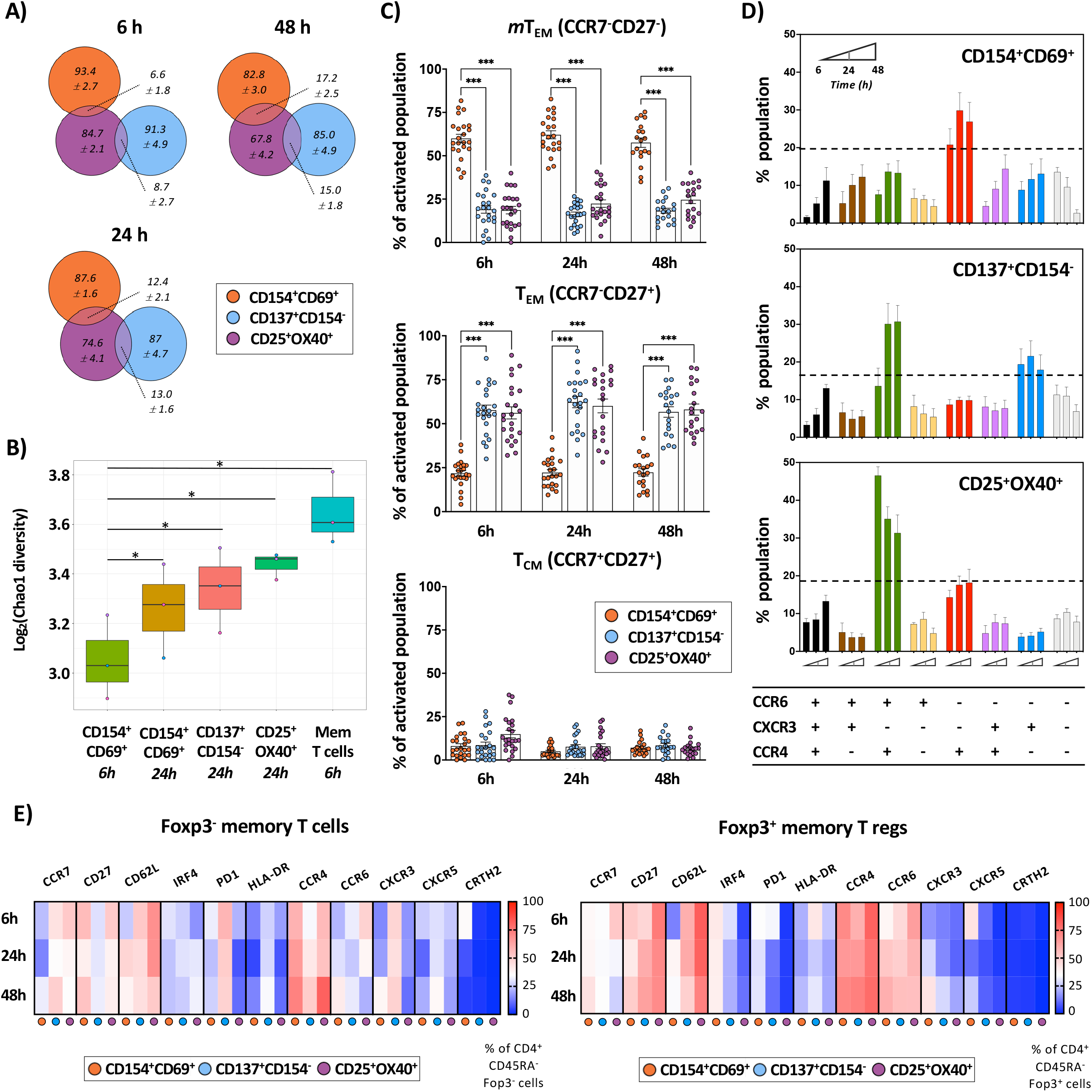
T cell responses to peanut allergens are associated with the activation of unique subsets of highly differentiated effector Th2 cells and memory regulatory T cells (Tregs) in peanut allergic (PA) subjects. **A**. Overlap of the activated populations after stimulation with crude peanut extract (CPE) for 6h, 24h, and 48h represented by Venn diagrams (n=24-30). Numbers inside the circles indicate the percentage (± SEM) of non-overlapped cells and numbers next to the lines specify the overlap percentage (± SEM). Different colors designate different activated populations. **B**. Estimated Chao1 alpha diversity of TCRβ repertoire in activated populations and total memory CD4^+^ T cells sorted from PA subjects stimulated with CPE for 6h and 24h (n=3). **C**. Percentage of the mature effector (*m*T_EM_), effector (T_EM_), and central (T_CM_) memory CD4^+^ T cells in the activated populations after stimulation of PBMCs from PA subjects with CPE for 6h, 24h, and 48h (n=25). **D**. Percentage of chemokine receptor co-expression in activated populations after CPE-stimulation for 6h, 24h, and 48h (n=5). **E**. Heatmaps of the percentage of expression for each marker in Foxp3^-^ T cells (left) and Tregs (Foxp3^+^, right) in activated populations after CPE-stimulation for 6h, 24h, and 48h (n=5). In (B) and (C) each data point is one individual (mean ± SEM) and bar graphs represent mean + SEM in (D). Statistical analyses by mixed-effect analysis with Geisser-Greenhouse correction followed by Tukey’s multiple comparisons test in (B) and (C). In (D), dashed line represents cut off above populations are significantly increased (mixed-effect analysis accounting for subject levels repeated measures at the three timepoints studied with Geisser-Greenhouse correction followed by Tukey’s multiple comparisons test, p<0.05. P values were corrected for false discovery rate). *P < 0.05, **P < 0.01, and ***P < 0.001.

We also studied the differentiation profile of the three distinct populations by dividing the memory T cell subsets into functional categories based on the expression of CCR7 and CD27 ^20,21^. Peanut-induced CD154^+^CD69^+^ T cells mainly lacked expression of CCR7 and CD27 (Fig. 3C), which is associated with terminally differentiated mature effector memory CD4^+^ T cells (*m*TEM). Significantly higher percentages of CD137^+^CD154^-^ and CD25^+^OX40^+^ T cells (p<0.001) were CCR7^-^ CD27^+^ effector memory CD4^+^ T cells (TEM) (Fig. 3C). Next, we analyzed chemokine receptor expression (CCR6, CXCR3, and CCR4) associated with distinct Th subsets ^22^. CD154^+^CD69^+^ T cells were enriched for CCR6^-^CXCR3^-^CCR4^+^ cells, consistent with their Th2 phenotype, whereas CD137^+^CD154^-^ T cells showed enrichment of CCR6^-^CXCR3^+^CCR4^-^ cells (consistent with a Th1 subset) and CCR6^+^CXCR3^-^CCR4^+^ (Fig. 3D). Interestingly, the CD25^+^OX40^+^ population was dominated by the expression of CCR4 and highly represented by CCR6^+^CXCR3^-^CCR4^+^ T cells (Fig. 2B). The expression of CCR6 and CCR4 in absence of CXCR3 has been associated with a Treg phenotype ^23^. To assess the characteristics of non-Tregs (Foxp3^-^) and Tregs (Foxp3^+^) in the activated populations, we performed spectral flow cytometry in the three peanut-induced T cell subsets. CD154^+^CD69^+^ cells mainly represented by Foxp3^-^ cells (Fig. 2D), showed an enriched expression of CRTH2 and PD1 at 6h that decreased at later timepoints (Fig 3E, left). Analysis of peanut-induced CD137^+^CD154^-^ T cells revealed two unique subsets contained within this population based on the expression of Foxp3 (Fig. 2D). Peanut-induced CD137^+^CD154^-^Foxp3^-^ T cells suggested an effector Th1 phenotype based on a high expression of CXCR3 and reduced expression of CCR4 and CCR6, which were characterized by an increased expression of PD1 and HLA-DR (Fig. 3E, left). By contrast, CD137^+^CD154^-^Foxp3^+^ T cells were characterized by an enriched expression of CCR4, CCR6, CCR7, and HLA-DR (Fig. 3E, right). Finally, CD25^+^OX40^+^ cells mainly represented by Foxp3^+^ cells (Fig. 2D), showed a high expression of CD27, CD62L, and CCR4, and a decreased expression of IRF4 and PD1 (Fig. 3E, right).

### Early secretion of IL-2 by peanut-specific CD154^+^CD69^+^ Th2 cells promotes activation of suppressive Tregs

To identify functional specialization of the peanut-induced T cell populations, we examined the production of Th2 (IL-5), Th1 (IFN-γ), and Treg (IL-10) cytokines by using cytokine secretion assays (Fig. 4A). The peanut-specific CD154^+^CD69^+^ population showed the highest secretion level of Th2 cytokines (IL-4, IL-5, and IL-13) during the first hours of CPE-stimulation (Fig. 4A; Fig. S4A) as well as IL-2 (Fig. S4A), which confirms the effector Th2 phenotype previously identified for this population (Fig. 3D). IFN-γ was singularly derived from CD137^+^CD154^-^ T cells at 6h of stimulation with CPE, as was the cytokine IL-10 at 2, 4, and 6h (Fig. 4A; Fig. S4A). These cells only produced some detectable levels of TNF-α after 6h stimulation (Fig. S4A). We further analyzed the functional profile of peanut-induced CD137^+^CD154^-^ T cells by intracellular cytokine staining using spectral flow cytometry. Results showed an increased expression of IL-2, IL-10, IFN-γ, and TNF-α (Fig. 4B), confirming our previous observation of two unique subsets within activated CD137^+^CD154^-^ T cells: a Th1-related subset (Fig. 3D) characterized by the expression of IFN-γ, TNF-α, and IL-2 (Fig. 4B), and a Treg-associated subset (Fig. 2D), with an increased expression of IL-10 (Fig. 4B). In contrast, no specific cytokine secretion was associated with the peanut-induced CD25^+^OX40^+^ T cells.

**Fig. 4.**
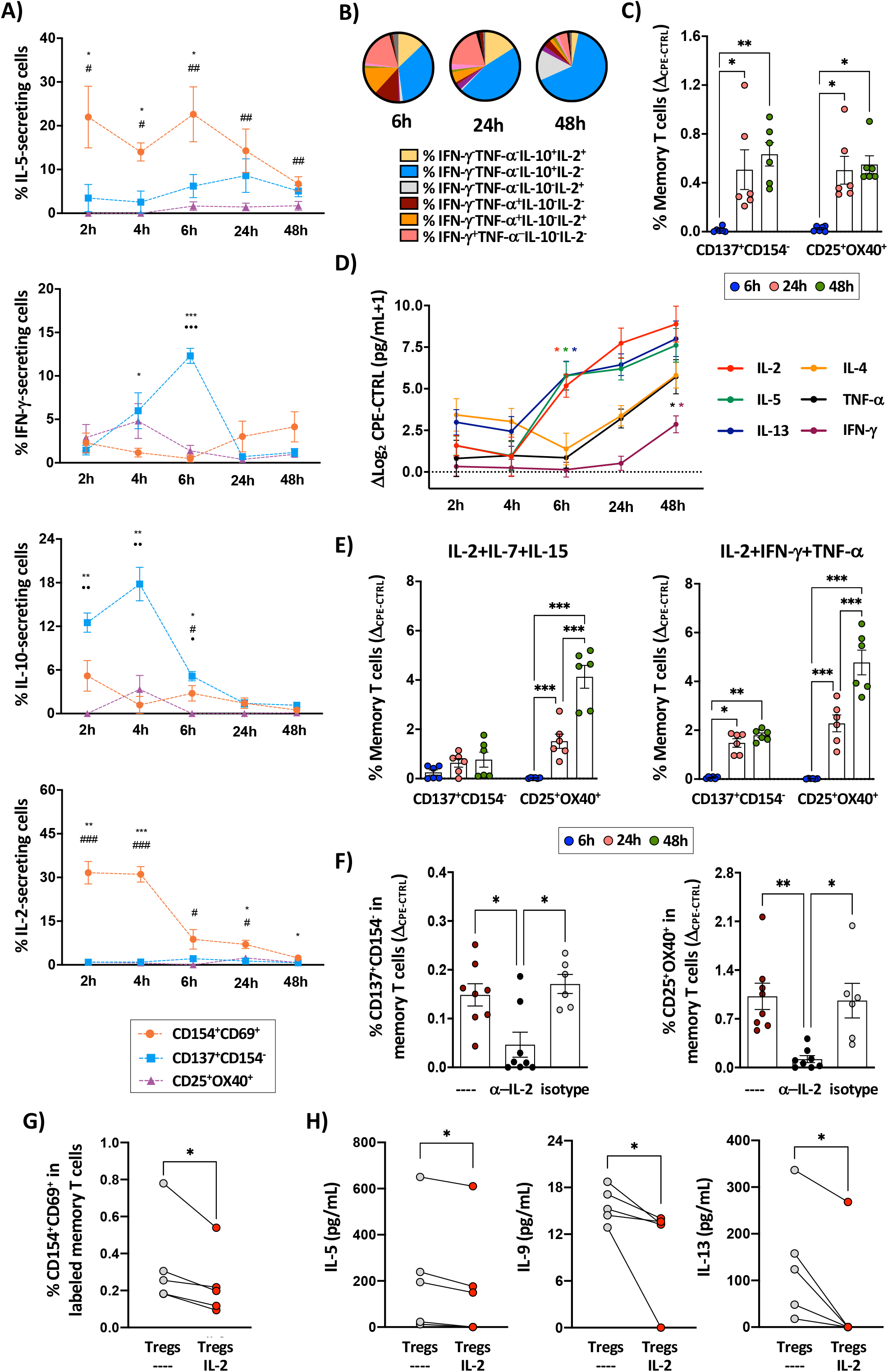
IL-2 released by the early activation of peanut-specific Th2 cells mediates the delayed upregulation of CD137, CD25, and OX40 on regulatory T cells (Tregs). **A**. Cytokine Secretion Assay (CSA) of activated populations after stimulation of PBMCs from peanut allergic (PA) subjects with crude peanut extract (CPE) for 2h-48h (n=6-11). **B**. Percentage of the intracellular cytokine co-expression for IFN-γ, TNF-α, IL-10, and IL-2 in CD137^+^CD154^-^ T cells after CPE-stimulation for 6h, 24h, and 48h (n=5). Non-cytokine-producing cells and populations ≤ 2.5% relative abundance are not represented. **C**. T cell activation induced after culture (24h) of PBMCs from non-allergic individuals with supernatants collected after CPE-stimulation of PBMCs from PA subjects for 6h, 24h, and 48h (n=6). **D**. Quantification of cytokines released to the culture supernatant by PBMCs from PA subjects stimulated with CPE for 2-48h (n=25-28). **E**. T cell activation induced by stimulation of PBMCs from non-allergic individuals with cytokine pools (IL-2+IL-7+IL-15, left; IL-2+IFN-γ+TNF-α, right) for 6h, 24h, and 48h. **F**. Neutralization of T cell activation by anti-IL-2 antibody and its isotype control in CPE-stimulated PBMCs from PA subjects for 24h (n=6-8). **G**. Percentage of CD154^+^CD69^+^ T cells in CellTrace-labeled PBMCs cultured with non-labeled purified Tregs alone or pre-incubated with IL-2 (24h) after CPE-stimulation for 6h (n=5). **H**. IL-5, IL-9, and IL-13 released by PBMCs cultured with purified Tregs alone or pre-incubated with IL-2 (24h) after CPE-stimulation for 6h (n=5). In (A) and (D) each point represents the mean ± SEM, one individual (mean ± SEM) in (C), (E), and (F), and one individual in (G) and (H). In (A), (C), (D), (E), and (F) statistical analysis by mixed-effect analysis with Geisser-Greenhouse correction followed by Tukey’s multiple comparisons test. In (A), * expresses differences between CD154^+^CD69^+^ and CD137^+^CD154^-^ cells, # expresses differences between CD154^+^CD69^+^ and CD25^+^OX40^+^ cells, and • expresses differences between CD137^+^CD154^-^ and CD25^+^OX40^+^ cells). In (D), * expresses the first significantly different timepoint (p<0.05) compared with baseline (2h) accounting for subject level repeated measure at the 5 different timepoints studies and each color refers its respective cytokine. In (G) and (H) statistical analysis by Wilcoxon’s test. *,#,•P<0.05; **,##,••P<0.01; and ***P<0.001.

We hypothesized that delayed activation of Treg subsets (CD137^+^CD154^-^ and CD25^+^OX40^+^) might be mediated by cytokines secreted by other cells. To test this, we applied the supernatants collected from PA cultures (at 6, 24, and 48h) to PBMCs from non-allergic donors. After 24h, we detected a significant increase in CD137^+^CD154^-^ and CD25^+^OX40^+^ activated T cells in non-allergic donors that had been incubated with the 24 and 48h supernatants from CPE-stimulated PA cultures (Fig. 4C), which suggests an effect of secreted cytokines on the induction of these activated populations. Next, we quantified the cytokines secreted to the cell culture supernatant by PBMCs from PA subjects after stimulation with CPE over time. Results revealed that IL-2 and the Th2 cytokines IL-5 and IL-13 were significantly increased as early as 6h after CPE-stimulation, and TNF-α and IFN-γ production enhanced at 48h (Fig. 4C). In order to identify the specific cytokines involved in the induction of CD137^+^CD154^-^ and CD25^+^OX40^+^, we evaluated the effects of three cytokine pools (IL-4+IL-5+IL-13, IL-2+IL-7+IL-15, and IL-2+TNF-α+IFN-γ) on PBMCs from non-allergic donors. Our results showed no significant impact of Th2 cytokines in any of the populations (Fig. S4B). By contrast, stimulation of non-allergic PBMCs with the pool of cytokines IL-2+IL-7+IL-15 induced CD25^+^OX40^+^ T cells after 24 and 48h, and the pool of IL-2+IFN-γ+TNF-α induced both populations (CD137^+^CD154^-^ and CD25^+^OX40^+^) after 24 and 48h of culture (Fig. 4E). Based on these results, we then neutralized cytokines in PBMCs from PA subjects during CPE-stimulation to evaluate the contribution of each of those cytokines to the specific induction of the T cell peanut-reactive populations (Fig. 4F; Fig. S4C and S4D). Together, neutralization assays consistently showed that IL-2 blockade suppressed the generation of CD137^+^CD154^-^ and CD25^+^OX40^+^ T cells.

To determine the regulatory capacity of IL-2-activated CD137^+^CD154^-^ and CD25^+^OX40^+^ T cells to control peanut-specific Th2 responses, we pre-incubated CD127_low_CD25^+^ Tregs from PA subjects with IL-2 for 24h prior to the specific stimulation of their PBMCs with CPE. Pre-incubation with IL-2 efficiently induced the generation of CD137^+^CD154^-^ and CD25^+^OX40^+^ Tregs (Fig. S4E) that were able to decrease the early induction of peanut-specific CD154^+^CD69^+^ Th2 cells (Fig. 4G). This reduction was also associated with a decreased secretion of the Th2 cytokines IL-5, IL-9, and IL-13 (Fig. 4H), while IL-2 production was preserved (Fig. S4F). Taken together, our data indicate that early production of IL-2 by peanut-specific CD154^+^CD69^+^ T cells from PA subjects induces an allergen-independent activation of two different Treg populations (CD137^+^CD154^-^ and CD25^+^OX40^+^) with a functional capacity to control peanut-induced Th2 allergic responses.

### In vivo exposure to peanut allergens results in a delayed and durable activation of Tregs characterized by their distinct upregulation of CD137 and OX40

Since a similar kinetics of activation *in vivo* might explain the expansion observed in CD127_low_CD25^+^ Tregs during egg OIT (Fig. 1A and 1B), we sought to validate the functional relevance of our *in vitro* findings by examining the dynamics of peanut-induced CD4^+^ T cell responses after a double-blind placebo-controlled peanut challenge (DBPCPC) in PA subjects. Firstly, we evaluated the CD4^+^ T cell activation in PBMCs collected from PA children before (baseline, T0) and 24h after a DBPCPC using spectral flow cytometry. As expected, due to the rapid degradation of CD154 (CD40L) following the interaction with the CD40 expressed in circulating B cells *in vivo*, we did not observe CD154 upregulation after DBPCPC. However, we did observe an increased expression of CD69 in pathogenic Th2A cells gated as CD4^+^CD45RA^-^CD27^-^ CD161^+^CRTH2^+^CD49d^+^ T cells (Fig. 5A). Consistent with an early activation of these allergen-specific Th2A cells *in vivo*, we observed a significant increase in plasma IL-5 levels 4h after the DBPCPC (p<0.01), and a delayed increase of IL-2 after 24h (p<0.001) (Fig. 5B). Although we did not find an expansion of memory CD127_low_CD25^+^ cells, when the IL-2Rα receptor (CD25) was evaluated in memory CD127^-^Foxp3^+^ T cells, a significant increase of this marker was detected 24h after the DBPCPC (p<0.05; Fig. 5C), suggesting that an IL-2-mediated secondary activation of Tregs could also be generated *in vivo*. In fact, this activation was confirmed by an increased expression of CD137, OX40, CD69, and PD1 in Tregs (CD25^+^CD127^-^Foxp3^+^) comparing baseline (T0) to 24h following the DBPCPC (Fig. 5D). We also observed a higher frequency of LAP^+^ Tregs 24h after the DBPCPC (Fig. 5D).

**Fig. 5.**
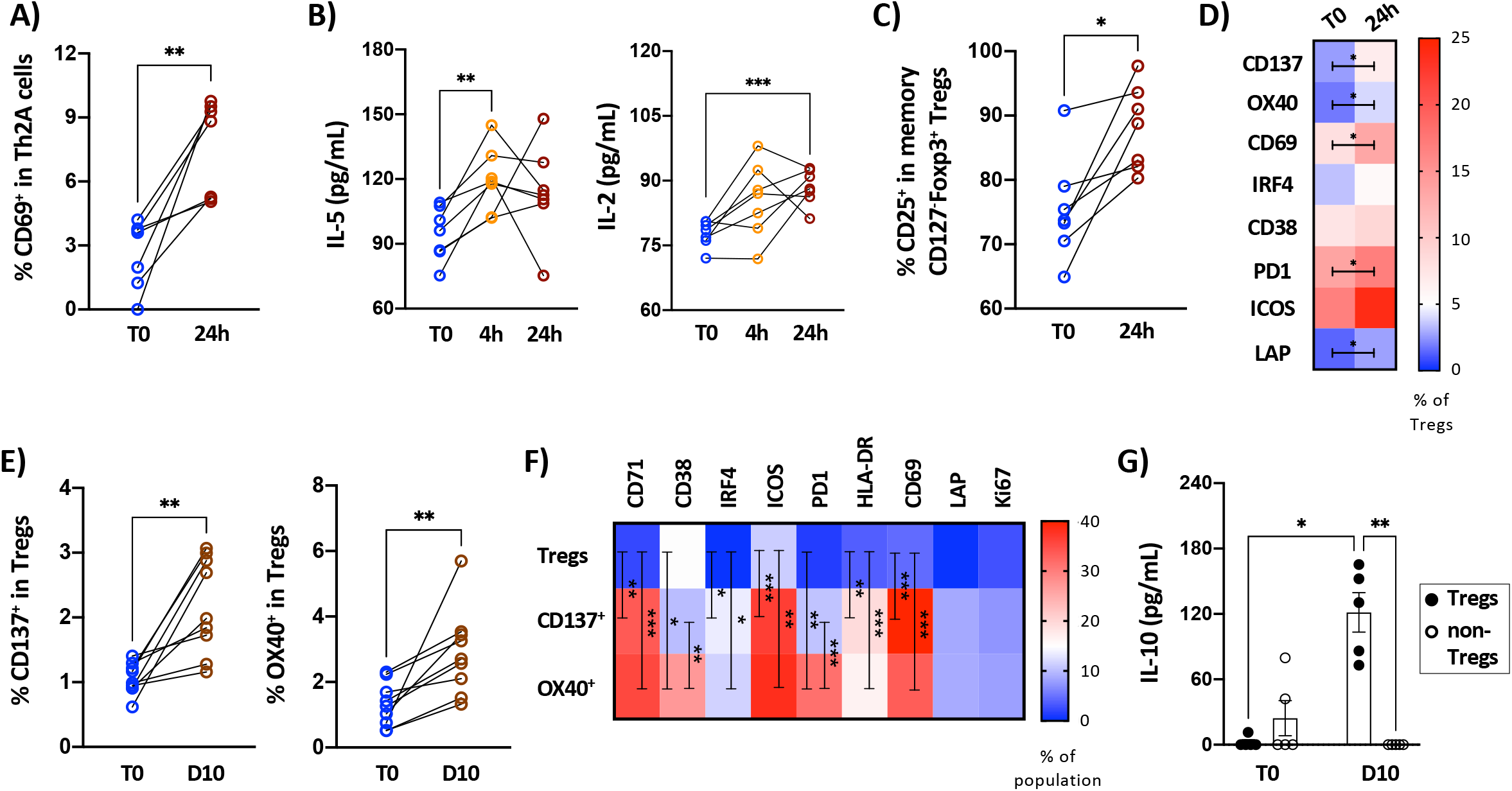
In vivo *recognition of peanut allergens generates an early activation of allergen-specific Th2 cells and a durable activation of regulatory T cells (Tregs) in peanut allergic (PA) subjects*. **A**. Percentage of CD69^+^ Th2A cells in PBMCs from PA subjects (n=7) before (T0) and after (24h) oral peanut challenge a double-blind placebo-controlled peanut challenge (DBPCPC). **B**. Quantification of IL-5 and IL-2 in sera from PA subjects before (T0) and after (4h and/or 24h) a DBPCPC (n=7). **C**. Percentage of CD25 expression in memory CD127^-^Foxp3^+^ T cells (n=7) before (T0) and after (24h) a DBPCPC (n=7). **D**. Heatmaps of the percentage of expression for each marker in Tregs (CD25^+^CD127^-^Foxp3^+^) before (T0) and after (24h) a DBPCPC (n=7). **E**. Percentage of CD137 and OX40 expression in memory Tregs (CD25^+^CD127^-^Foxp3^+^) before (T0) and after (10d) a DBPCPC (n=9). **F**. Heatmaps of the percentage of expression for each marker in Tregs (CD25^+^CD127^-^Foxp3^+^), CD137+ Tregs, and OX40+ Tregs 10d after a DBPCPC (n=9). **G**. IL-10 released by PBMCs from PA subjects obtained before (T0) and after (10d) a DBPCPC (n=5) and CPE-stimulated for 72h with and without depletion of Tregs (CD127_low_CD25^+^). In (A), (B), (C), (E), and (G), each data point is one individual. In (A), (C), and (E) statistical analysis by paired Student’s t test and mixed-effect analysis with Geisser-Greenhouse correction followed by Tukey’s multiple comparisons test is used in (B), (D), and (F). In (G) statistical analysis by Kruskal-Wallis test followed by Dunn’s multiple comparisons test. *P<0.05, **P<0.01, and ***P<0.001.

To establish whether the functional activation of CD4^+^ T cells was preserved over time *in vivo*, we studied their activation in PBMCs from PA individuals before (baseline, T0) and 10d after the DBPCPC. We found that Th2A cells were no longer activated 10d after the DBPCPC based on their expression of CD69 (Fig. S5B). In addition, no expansion of memory CD127_low_CD25^+^ T cells or increased expression of CD25 within CD127^-^Foxp3^+^ were observed (Fig. S5C and S5D). However, our results revealed a significantly increased upregulation (p<0.01) of the activation markers CD137 and OX40 in Tregs (CD25^+^CD127^-^Foxp3^+^) 10d after the DBPCPC, consistent with an efficient and durable activation of Tregs after *in vivo* recognition of peanut allergens in PA subjects (Fig. 5E). Both CD137^+^ and OX40^+^ activated Tregs showed an increased expression of CD71, IRF4, ICOS, HLA-DR, and CD69 compared with total memory CD25^+^CD127^-^Foxp3^+^ Tregs (Fig. 5F). OX40^+^ cells exhibited a unique upregulation of CD38 and PD1 when compared with CD137^+^ and total memory Tregs (Fig. 5F), supporting a long-term activation of this population *in vivo*. Finally, in order to evaluate the functionality of this durable Treg activation, we depleted Tregs from PBMCs of PA individuals who followed the DBPCPC. Our results showed a significantly increased production of the regulatory cytokine IL-10 following 10d of the DBPCPC (p<0.05) that was abolished when Tregs were depleted (Fig. 5G), suggesting the *in vivo* generation of IL-10-producing Tregs after oral exposure to peanut in PA subjects. Taken together, these results suggested that early activation of allergen-specific peTh2 cells *in vivo* could also induce an IL-2-mediated durable activation of Tregs that may contribute to the induction of a sustained oral tolerance to food allergens via IL-10 secretion.

## DISCUSSION

In this work we have investigated the kinetics of activation and functional characteristics of food allergen-induced CD4^+^ T cells from peanut allergic children activated *in vitro* and *in vivo*. Although type 2 CD4^+^ T cells of various phenotypes have been described (Th2A, peTh2, and Tfh13), other food allergen-specific T cell subsets such as Tregs and their relationship with effector Th2 cells remain poorly understood ^3,7,24^. We observed an expansion of total Tregs (CD127_low_CD25^+^) in egg allergic children with positive clinical response after receiving OIT. Despite highlighting the importance of Tregs in the clinical response, the allergen specificity of these Tregs was not clear. We used a cohort of PA subjects to evaluate the expression of activation markers in their PBMCs stimulated with peanut at different timepoints. Our data revealed three unique populations of peanut-induced memory CD4^+^ T cells based on distinct activation patterns (CD154^+^CD69^+^, CD137^+^CD154^-^, and CD25^+^OX40^+^) upon allergen recognition. An early activation (6h) with a restricted TCRβ clonal diversity was found in CD154^+^CD69^+^ T cells, consistent with a highly peanut-specific clonotype-driven response ^7,25,26^. We also observed a delayed activation of CD4^+^ T cells expressing CD137 and CD25/OX40. These latter memory T cell populations had diverse TCRβ repertoires with minimal overlap with the repertoire of CD154+ cells.

The characteristics of the cells contained within the CD154^+^CD69^+^ population are consistent with these cells being effector type 2 cells as previously described in peanut allergy ^3,4^. Specifically, CD154^+^CD69^+^ T cells were characterized by a highly mature profile of differentiation (CD27^-^CCR7^-^), a Th2 surface marker phenotype (CCR6^-^CXCR3^-^CCR4^+^), and characterized by very rapid (<6h) secretion of multiple Th2-related (IL-4, IL-5, and IL-13) and effector (IL-2 and TNF-α) cytokines upon allergen recognition, all consistent with previously described peanut-specific peTh2 cells ^3,5,7^. Functionally, we showed that CD154^+^CD69^+^ T cells have all the necessary components, including CD40L expression, IL-4, and homing molecules such as CCR6 or CXCR5 to promote IgE class-switching at effector sites including the gastrointestinal tract or in the B cell follicles. Finally, we found that a single oral exposure to peanut *in vivo* in the form of an oral challenge induces the activation of Th2A cells (CD69^+^) and the systemic release of the type 2 cytokine IL-5 and IL-2 in PA individuals.

The second function observed for CD154^+^CD69^+^ peanut-specific peTh2 cells was the induction of a second wave of T cell activation involving Treg populations via IL-2 secretion. CD137^+^CD154^-^ and CD25^+^OX40^+^ peanut-responsive T cells were dependent on IL-2 for their activation, and unlike CD154^+^CD69^+^ T cells, showed no evidence of reduced TCRβ clonal diversity. It has been described that once allergen is recognized by specific CD4^+^ T cells, a bystander amplified response is initiated by interaction of IL-2 with its receptors (including IL-2Rα, encoded by *IL2RA*, and also known as CD25) playing singular roles in different T cell subsets and prompting a strong activation of Tregs ^27,28^. Tregs do not produce IL-2 and rely on the paracrine production of IL-2 by activated effector T cells. *In vivo* results have revealed that paracrine IL-2 production by antigen-specific T cells plays a critical role in the activation of STAT5 and initiates a local negative feedback based on the increased suppressive activity of Tregs ^28^. This paracrine IL-2-mediated feedback that enhances suppressive functions and proliferation of co-localized Treg has been reported to rapidly constrain antigen-specific effector CD4+ T cells, ultimately limiting their division and inducing their apoptosis ^29^. We found a persistent activation of Tregs after allergen exposure *in vivo* characterized by the distinct upregulation of CD137 and OX40, and the expression of immunoregulatory molecules, including LAP (a component of surface TGF-β), PD-1, and ICOS, that contributed to an enhanced production of IL-10. CD137 and OX40 are members of the tumor necrosis factor receptors superfamily (TNFRSF) along with others such as CD40 and they act as T cell co-stimulatory molecules being involved in potentiating T cell responses triggered through TCR engagement. Their ligands CD137L (4-1BBL) and OX40L are expressed by antigen presenting cells (APCs). Thus, antigen stimulation and subsequent signaling through the TCR results in CD137 and OX40 upregulation in effector T cells and Tregs ^30-32^. However, a TCR-independent induction of CD137 and OX40 driven by IL-2 has been also reported in Tregs, associating this cytokine-mediated upregulation to the maturation of Tregs ^33-35^. Together, CD137^+^ and OX40^+^ activated Tregs could act back on Th2 cells via APCs or act directly on allergic effector cells such as mast cells to suppress immediate hypersensitivity.

In the context of immunotherapies for the treatment of airborne allergens in humans, there is evidence for an increased proportion of peripheral antigen-specific Tregs ^36^ and this has formed the framework for our understanding of the mechanism of immunotherapy, despite the fact that there is no conclusive evidence of the expansion of food allergen-specific Tregs after a successful OIT. When peanut-specific CD4^+^ T cells were studied using tetramers before and after allergen OIT, an increase in frequency of anergic T cells and a decrease in peTh2 cells was found, but no increase in peanut-specific Tregs was observed ^24^. Monian et al. recently used single cell RNAseq of peanut-responsive (CD154^+^ or CD137^+^) cells captured 20h after peanut stimulation ^37^. They did not observe a significant difference in Treg phenotype after OIT, an induction of new clonotypes within the Treg compartments, or a decreased frequency of peanut-specific CD137^+^ cells after OIT. Indeed, reports of increased Tregs after food OIT have either used proliferation-based approaches, which do not discriminate between TCR or cytokine-induced activation, or have quantified total Tregs ^10,38-40^. We found that beyond the previously-reported reduction of allergen-specific Type 2 cells in egg allergic children receiving allergen OIT ^41^, a significant expansion of total Tregs (CD127_low_CD25^+^) was observed. Furthermore, sub-group analysis demonstrated that Treg expansion was only observed in those with the greatest clinical benefit. Our work is consistent with that reported by Karlsson et al. on the natural outgrowth of cow’s milk allergy ^42^. They observed that dietary milk re-introduction to tolerant children, but not allergic children, led to an expansion of circulating CD4^+^CD25^+^ Tregs with the ability to suppress effector T cell responses to milk allergens. There is an intriguing disconnect between the impact of OIT on total Treg populations, which increase, and those described as allergen-specific, which are unchanged. The clinical response to allergen immunotherapy is allergen-specific, indicating that protective mechanisms must also be allergen-specific. Our data indicate that Treg specificity of action is derived in part from the effector cell release of IL-2, and when OIT is stopped, Treg populations begin to contract. This is in agreement with the POISED trial that. reported a progressive loss of tolerance over time with sustained peanut avoidance after allergen OIT ^43^, which suggests that continuous peanut exposure is needed to maintain long-term tolerance. Administration of IL-2 has been tested for the preferential expansion of suppressive Tregs as a part of immunotherapies in many inflammatory and autoimmune diseases using two different strategies: i) IL-2/α-IL-2 complex for the treatment of inflammatory colitis ^44^ and type 1 diabetes ^45^; ii) low doses of IL-2 to enhance the numbers of Tregs in graft-versus-host disease ^46^, type 1 diabetes ^47^, hepatitis C virus-induced vasculitis ^48^, and systemic lupus erythematosus ^49^. In addition, recent studies in animal models of food allergy and allergic asthma have reported that both IL-2-based therapeutic approaches are able to efficiently induce tolerance to these antigens ^50,51^. We hypothesize that administration of IL-2 to selectively expand Tregs could be of utility in the treatment of food allergy.

In conclusion, we have determined that there are two unique waves of T cell activation in response to food allergens in allergic individuals, an initial allergen-specific effector type 2 response and a delayed IL-2-dependent activation of Tregs. These latter cells have a suppressive ability *in vitro*, are induced *in vivo* after a single oral exposure to the food allergen, and are associated with a sustained activation. Together, we speculate that IL-2-mediated activation of Tregs is an important mechanism for the restoration oral tolerance.

## MATERIALS AND METHODS

### 1. Participant recruitment and blood processing

Peanut allergic (PA) subjects and egg allergic individuals were recruited at Jaffe Food Allergy Institute and pediatric non-allergic donors consented at Susan and Leonard Feinstein IBD Center (both at Mount Sinai Hospital, New York, NY). Clinical information obtained from these participants is summarized in Table S1. For PA subjects undergoing double-blind placebo-controlled peanut challenge (DBPCPC) included in this study (Table S1), blood samples were collected at baseline avoiding peanut before DBPCPC (T0) as well as 4h, 24h and 10d after DBPCPC with peanut (≥ 143mg of protein). Informed consents were obtained from all participants or their parents/guardians following the protocols approved by the Institutional Review Board of the Icahn School of Medicine at Mount Sinai. All peripheral blood samples were collected in sodium-heparin vacutainer tubes, IgE specific antibodies for peanut and egg proteins measured by ImmunoCAP (Thermo Fisher Scientific, Waltham, MA) in plasma samples, and PBMCs isolated by density gradient using Ficoll-Paque PLUS (GE Healthcare, Chicago, IL) and cryopreserved in AB serum (GemCell, Gemini-Bioproducts, West Sacramento, CA) containing 10% DMSO (Sigma-Aldrich, St Louis, MO) in liquid nitrogen until further use. Three buffy coats were obtained from the New York Blood Center (New York, NY) and PBMCs were isolated and preserved as indicated above. Finally, egg allergic subjects following oral immunotherapy (OIT) with raw egg white were enrolled in the multicenter Consortium of Food Allergy Research (COFAR) intervention (COFAR7, NCT01846208) of which clinical details, as well as PBMC processing and analysis protocols used in this clinical trial, were previously published ^9,18,41^.

### 2. PBMC stimulation and cytokine quantification

Cryopreserved PBMCs were thawed, washed, plated in 24-well plates at a concentration of 4 × 10^6^ cells/mL in 1 mL of AIM V medium (Gibco, Waltham, MA) containing 2.5% of AB serum (GemCell), and rested overnight (ON). The day after, cells were unstimulated (control [CTRL]), stimulated with 100 µg of protein/mL (crude peanut extract [CPE] and egg white [EW]), or 1.25 µL of anti-CD3/CD28 stimulation beads (Thermo Fisher Scientific) for a bead:cell ratio of 1:5. For surface CD154 detection, 1µg/mL of the blocking anti-human CD40 antibody (clone HB14; BioLegend, San Diego, CA) was added to maintain CD154 at the cellular surface and PBMCs were cultured under standard conditions (5% CO_2_ and 37°C) for 2, 4, 6, 24 and/or 48h. Both CPE and EW were cleaned of endotoxins by using Detoxi-Gel columns (Thermo Fisher Scientific), residual endotoxin levels quantified with a LAL assay (Thermo Scientific, Waltham, MA) and reduced below 0.1 EU/mL (99.85% removal and within the acceptable range for culture reagents). After culture, cells were harvested, stained for their phenotypical analysis by flow cytometry using different flow panels that combined the antibodies listed in Table S2, and acquired in a LSR Fortessa cytometer (BD Biosciencess, Franklin Lakes, NJ). For the high-dimensional characterization of memory CD4^+^ T cells in PA patients and PA individuals undergoing a DBPCPC by spectral flow cytometry, cells were labeled with surface and intracellular markers (Table S3 and S4) and analyzed in Cytek™ 4-laser and 5-laser Aurora instruments (Cytek Biosciences, Fremont, CA). In some experiments, supernatants were collected after culture and cytokines quantified using a human Th cytokine 13-multiplex assay (IL-1β, IL-13, IL-33, IL-2, IL-4, IL-3, GM-CSF, TNF-α, IL-6, IL-5, IL-15, IL-21, and IL-7; LEGENDPlex™, BioLegend) following manufacturer’s instructions. Samples were acquired in a CytoFLEX device (Beckman Coulter, Jersey City, NJ) and data analyzed with the LEGENDplex™ Data Analysis Software Suite (BioLegend).

### 3. Immunosuppression assay and regulation of Th2 cytokine secretion by Tregs

Cryopreserved PBMCs isolated from buffy coats of healthy donors (n=3) were thawed, washed, rested ON, and stimulated with 1.25 µL of anti-human CD3/CD28 stimulation beads (bead:cell ratio of 1:5; Thermo Fisher Scientific) for 24 h. After incubation, memory CD4^+^ T cells were negatively enriched using an EasySep™ Human Memory CD4+ T Cell Kit (STEMCELL Technologies, Kent, WA) and stained with a flow cytometry panel of surface markers (Table S5). Activated Treg populations (CD137^+^CD154^-^ and CD25^+^OX40^+^ T cells) were FACS-sorted following the gating strategy shown in Fig. S2B using a FACS Aria II instrument (BD Biosciences). In addition, responder CD25^-^CD4^+^ T cells (Tresps) were negatively purified from the autologous PBMC sample by using EasySep™ Human CD4^+^ T Cell isolation and Pan-CD25 depletion kits (STEMCELL Technologies). Tresps were CFSE-labeled (CellTrace CFSE, Thermo Fisher Scientific) following manufacturer’s protocol, and a total of 1 × 10^5^ Tresps were co-cultured in different ratios (Treg:Tresp, 1:1, 1:5, and 1:10) with the autologous FACS-purified activated Treg population in 96-well U-bottom plates. Co-cultures were polyclonally stimulated with anti-human CD3/CD28 beads (bead:cell ratio of 1:5; Thermo Fisher Scientific) under standard conditions (5% CO2 and 37°C) for 5d. For negative suppression control wells, Tresps were added alone with polyclonal stimulation, whereas negative proliferation control wells were prepared with Tresps in absence of anti-human CD3/CD28 beads. After 5d, T cells were harvested, stained for flow cytometry analysis using the panel described in Table S6, and analyzed on a CytoFLEX instrument (Beckman Coulter). The percentage of inhibition of CFSE-labeled Tresps was analyzed using Cytobank software (Mountain View, CA) following the formula: % inhibition= ([proliferated Tresps in negative suppression control – proliferated Tresps in ratio 1:X / total proliferated Tresps] x 100). All suppression experiments were performed in triplicate.

Regulation of Th2 cytokine secretion by activated Tregs was evaluated using Th2-polarized cells co-cultured with CD137^+^CD154^-^ and CD25^+^OX40^+^ activated T cells. Th2 cells were polarized by stimulating PBMCs isolated from buffy coats of healthy donors (n=3) with 1.25 µL of anti-human CD3/CD28 stimulation beads (bead:cell ratio of 1:5; Thermo Fisher Scientific) and IL-4 (20 ng/mL; Peprotech, Rocky Hill, NJ) in presence of anti-human IFN-γ (10 µg/mL; clone B133.5; BioXcell, Lebanon, NH) and IL-12p70 antibodies (10 µg/mL; clone 20C2; BioXcell) for 8d. After polarization, CD4+ T cells were enriched using EasySep™ Human CD4+ T Cell Kit (STEMCELL Technologies). CD137^+^CD154^-^ and CD25^+^OX40^+^ T cells were FACS-sorted from PBMCs stimulated with 1.25 µL of anti-human CD3/CD28 stimulation beads (bead:cell ratio of 1:5; Thermo Fisher Scientific) for 24 h as described above (Fig. S2B; Table S5). A total of 1 × 10^6^ Th2-polarized cells were co-cultured in different ratios (Treg:Th2, 0:1, 1:1, and 1:10) with the autologous FACS-purified activated T cell population in 24-well plates and stimulated with 1.25 µL of anti-human CD3/CD28 stimulation beads (bead:cell ratio of 1:5; Thermo Fisher Scientific) for 72h. Supernatants were collected after culture and cytokines quantified using a human Th cytokine 13-multiplex assay (IL-1β, IL-13, IL-33, IL-2, IL-4, IL-3, GM-CSF, TNF-α, IL-6, IL-5, IL-15, IL-21, and IL-7; LEGENDPlex™, BioLegend) following manufacturer’s instructions. Samples were acquired in a CytoFLEX device (Beckman Coulter) and data analyzed with the LEGENDplex™ Data Analysis Software Suite (BioLegend).

### 4. TCR sequencing and data analysis

After stimulation of PBMCs from PA subjects with CPE for 6 and 24h, memory CD4+ T cells were negatively enriched using an EasySep™ Human Memory CD4^+^ T Cell Kit (STEMCELL Technologies) and stained with a flow cytometry panel of surface markers (Table S5). Activated populations (CD154^+^CD69^+^ at 6 and 24h; CD137^+^CD154^-^ at 24h; and CD25^+^OX40^+^ at 24h) as well as total memory T cells (CD45RA^-^CD4^+^ T cells at 6h) were FACs-sorted by using a FACS Aria II device (Fig. S2B; BD Biosciences). Sorted T cells were lysed in Buffer RLT Plus (Qiagen, Hilden, Germany) and stored at -80°C until isolation of the genomic DNA (gDNA) using the QIAamp DNA Micro Kit (Qiagen) following manufacturer’s instructions. Sequencing of the complementarity-determining region 3 (CDR3) of human TCRβ chains and computational identification of clones was performed by immunoSEQ assay via Adaptive Biotech. (Adaptive Biotechnologies, Seattle, WA), as previously described ^52^. Briefly, gDNA was amplified in a bias-controlled multiplex PCR, followed by high-throughput sequencing by Illumina NextSeq platform. Raw sequence reads were demultiplexed according to Adaptive Biotech. proprietary barcode sequences. Demultiplexed reads were then further processed to remove adapter and primer sequences; identify and remove primer dimer, germline and other contaminant sequences. The filtered data was clustered using both the relative frequency ratio between similar clones and a modified nearest-neighbor algorithm, to merge closely related sequences to correct for technical errors introduced through PCR and sequencing. The resulting sequences were sufficient to allow annotation of the V, D, and J genes and the N1, N2 regions constituting each unique CDR3 and the translation of the encoded CDR3 amino acid sequence. Gene definitions were based on annotation in accordance with the IMGT database (www.imgt.org). The set of observed biological TCRβ CDR3 sequences were normalized to correct for residual multiplex PCR amplification bias and quantified against a set of synthetic TCRβ CDR3 sequence analogues. Data was analyzed using the immunoSEQ Analyzer toolset ^52-54^. TCR sequences were analyzed as a single batch, mitigating any potential confounding batch effects.

#### 4.1. TCR data normalization via downsampling

TCR diversity and clonotype analysis was performed using the R package called immunarch ^55^. Because of a large range in total clones sequenced by sample, for diversity measures we employed a downsampling approach that simulates all clones within a subject, detects the flow-sorted population within the subject that contains the lowest number of sequenced clones; all other flow-sorted populations for that subject are randomly sampled to an equivalent depth as the population with the lowest number of clones. In this way, each individual’s cellular populations are sampled to an equal depth, thus removing the confounding effects of differential clone detection.

#### 4.2. Accounting for different clone depth per individual

Given the above described downsampling procedure, there is a subject level technical effect of depth. Within each subject, all sample types (CD154^+^CD69^+^, CD137^+^CD154^-^, CD25^+^OX40^+^, and total memory CD45RA^-^ cells) are held to a constant clonal depth. However, across subjects, there is a difference that is limited by that particular subject’s population with the lowest clonal depth. To account for this across subject difference in depth, subject was used as a fixed effect covariate for all statistical comparisons. Note that because there was only a single measure per subject:sample-type pair there were not “repeated-measures” – but rather only a single measure, with a paired covariate across sample-types: subject.

### 5. Cytokine secretion assays and intracellular cytokine staining

For the identification of live cytokine-producing CD4^+^ T cells, we used a cytokine secretion assay (CSA; Miltenyi Biotec, Bergisch Gladbach, Germany) based on the capture of secreted cytokines by a cell surface-bound capture antibody followed by detection with a fluorescent anti-cytokine antibody. Briefly, cryopreserved PBMCs from PA individuals were thawed, seeded in a 24-well plate at 4 × 10^6^ cells/mL, rested ON, and stimulated with CPE (100µg of protein/mL) for 2h, 4h, 6h, 24h and 48h in presence of a blocking anti-human CD40 antibody (1µg/mL; clone HB14; BioLegend) for the detection of cell surface CD154. After incubation time, cells were harvested, washed with ice-cold working buffer (0.5% BSA, 2mM EDTA in PBS), and labeled with capture antibodies for IL-2, IL-4, IL-5, IL-10, IL-13, TNF-α, and IFN-γ (Miltenyi Biotec) in AIM V medium (Gibco) supplemented with 2.5% AB serum (GemCell) at 4°C for 5 min, following manufacturer’s instructions. Cells were further diluted in pre-warmed medium (2.5% AB serum in AIM V) at a concentration of 1 × 10^6^ cells/mL and incubated at 37°C under rotation (100 rpm) for 45 min. Finally, cells were washed with working buffer (0.5% BSA, 2mM EDTA in PBS), stained for viability and extracellular markers (Table S7), and analyzed by conventional flow cytometry in a LSR Fortessa instrument (BD Biosciencess).

The intracellular cytokine staining (ICS) experiments were carried out with cryopreserved PBMCs from PA subjects that were similarly cultured and stimulated with CPE (100 µg of protein/mL) for 6h, 24h, and 48h in absence of blocking anti-human CD40 antibody. For the cytoplasmic detection of CD154 and the ICS, GolgiPlug (1µg/mL; BD Biosciencess) was added to the culture 4h before harvesting the cells. Surface and intracellular staining for spectral flow cytometry analysis was performed according to Table S8 and samples were analyzed in a Cytek™ 4-laser Aurora cytometer (Cytek Biosciences).

### 6. Induction and neutralization of bystander activated T cell populations

Supernatants collected from PA subject PBMCs (n=6) stimulated with CPE for 6, 24, and 48h (0.5 mL) were applied over 2 × 10^6^ PBMCs isolated from buffy coats of non-allergic individuals (n=3; New York Blood Center) in presence of an anti-human CD40 antibody (1 µg/mL; clone HB14; BioLegend) for 24 h. In addition, PBMCs from non-allergic patients were cultured with three different pools of cytokines for 6, 24, and 48h: i) IL-4 (50 IU/mL) + IL-5 (60 IU/mL) + IL-13 (10 IU/mL); ii) IL-2 (100 IU/mL) + IL-7 (20 IU/mL) + IL-15 (20 IU/mL); iii) IL-2 (100 IU/mL) + TNF-α (200 IU/mL) + IFN-γ (200 IU/mL); all from Peprotech. Finally, PBMCs from PA subjects (n=6) were stimulated with CPE (100 µg of protein/mL) alone or in presence of three pools of blocking anti-human antibodies (2 µg/mL) for 24 h: i) IL-4 (clone MP4-25D2, Invitrogen) + IL-5 (clone TRFK5, Invitrogen) + IL-13 (clone PVM13-1, Invitrogen); ii) IL-2 (clone AB12-3G4, Invitrogen) + IL-7 (clone BVD10-40F6, BioLegend) + IL-15 (clone ct2nu, Invitrogen); iii) IL-2 (clone AB12-3G4, Invitrogen) + TNF-α (clone MAb1, Invitrogen) + IFN-γ (clone NIB42, Invitrogen). Similarly, individual neutralization experiments were performed in presence of blocking anti-human IL-2, IL-15, TNF-α, IFN-γ (2 µg/mL; clones detailed above and all from Invitrogen), and IL-7 (2 µg/mL; clone BVD10-40F6, BioLegend) antibodies. Before stimulation, ON rested PBMCs were pre-incubated with the antibodies (or their pools) for 1h prior to adding CPE. Corresponding isotype antibodies were used as negative controls in these experiments (2 µg/mL; all from Invitrogen; Rat IgG1 kappa isotype control from BioLegend). In all the experiments described above, cells were harvested after incubation time and stained for the identification of the activated populations CD137^+^CD154^-^ and CD25^+^OX40^+^ within memory CD4^+^ T cells by flow cytometry (Table S9) using a LRS Fortessa device (BD Biosciences).

### 7. Functional activation of Tregs by IL-2 and Treg depletion

Cryopreserved PBMCs from PA subjects (n=5) were thawed, washed, rested ON, and Tregs purified by using EasySep™ Human CD4^+^CD127_low_CD25^+^ Regulatory T Cell Isolation Kit (STEMCELL Technologies). Remaining PBMCs (Treg-depleted) were labeled with CellTrace™ Violet Cell Proliferation Kit (Thermo Fisher Scientific) following manufacturer’s protocol. Purified Tregs were either unstimulated or stimulated with 5 ng/mL of IL-2 (Peprotech) for 24h. After incubation, a total of 8 × 10^4^ Tregs (unstimulated and IL-2-stimulated) were washed and co-cultured with 4 × 10^6^ CellTrace-labeled PBMCs in a 24-well plate and stimulated with CPE (100µg of protein/mL) for 6h in presence of a blocking anti-human CD40 antibody (1µg/mL; clone HB14; BioLegend) for the detection of cell surface CD154. Supernatants were collected and stored at - 80°C until further use and cells were harvested, stained for surface and intracellular markers (Table S10), and analyzed in a Cytek™ 5-laser Aurora cytometer (Cytek Biosciences). Finally, cryopreserved supernatants were used for the quantification of the cytokine secreted by using a human Th cytokine 13-multiplex (IL-5, IL-13, IL-2, IL-6, IL-9, IL-10, IFN-γ, TNF-α, IL-17A, IL-17F, IL-4, IL-21, and IL-22; LEGENDPlex, BioLegend) following manufacturer’s instructions. Samples were acquired on a CytoFLEX cytometer (Beckman Coulter) and raw data were analyzed with the LEGENDplex™ Data Analysis Software Suite (BioLegend).

PBMCs from PA subjects who underwent a DBPCPC (n=5) were thawed, washed, rested ON, and stained with a flow cytometry panel of surface markers (Table S11). CD127_low_CD25^+^ Tregs were sorted out by using a FACS Aria II device (BD Biosciences) and 4 × 10^6^ Treg-depleted PBMCs were cultured in a 24-well plate and stimulated with CPE (100µg of protein/mL). Non-Treg-depleted PBMCs were used as control. After 72h, supernatants were collected and used for the quantification of the cytokine secreted by using a human Th cytokine 13-multiplex (IL-5, IL-13, IL-2, IL-6, IL-9, IL-10, IFN-γ, TNF-α, IL-17A, IL-17F, IL-4, IL-21, and IL-22; LEGENDPlex, BioLegend) following manufacturer’s instructions. Samples were acquired on a CytoFLEX cytometer (Beckman Coulter) and raw data were analyzed with the LEGENDplex™ Data Analysis Software Suite (BioLegend).

### 8. Flow cytometry and data analysis

For the different flow cytometry staining performed in this study, harvested cells were firstly labeled for viability (Live/Dead Fixable dyes, Invitrogen and BioLegend), and washed in FACS buffer (2% FBS, 2mM EDTA in PBS). Fc receptors were blocked with Human TruStainFcX (BioLegend). Staining of cell surface markers was performed on ice for 30-40 min (Tables S2-S11). For combined intranuclear staining, cells were fixed/permeabilized by using the FoxP3/Transcription Factor Staining Buffer Set (Invitrogen) following manufacture’s recommendations and stained with intracellular antibodies (Tables S2-4, S6, and S9-10) on ice for 30-40 min. For a combined intracellular CD154 detection and ICS, cells were washed in FACS buffer, fixed in a 4% paraformaldehyde solution (Electron Microscopy Sciences, Hatfield, PA) at RT for 10 min, treated with permeabilization buffer (Invitrogen) at RT for 20 min, and stained with labeled antibodies (Table S8) on ice for 30 min. Stained cells were subsequently analyzed by using a CytoFLEX (Beckman Coulter), a LSR Fortessa™ (BD Biosciences), or Cytek™ 4-laser and 5-laser Aurora (Cytek Biosciences) cytometers. All samples were analyzed by using Cytobank. platform (Mountain View, CA) or FlowJo v 10.6.0 software (Tree Star Inc., Ashland, OR).

On the other hand, for the high-dimensional phenotypical and functional characterization of the memory CD4^+^ T cells of PA subjects by spectral flow cytometry (Table S3), unstimulated and CPE-stimulated samples for 6, 24, and 48 h were analyzed by an unsupervised computational analysis using FlowSOM algorithm ^56^, which uses a self-organizing map (SOM) followed by consensus hierarchical clustering to detect cell populations. After a manual gating to remove debris, doublets, and to select live memory (CD45RA^-^) CD4^+^CD3^+^ T cells, a proportional sampling of 10,000 events was randomly applied across samples and the marker expression values were arcsinh-transformed with a co-factor of 5 operating a pre-determined number of 9 clusters. To train the SOM, 1,000 interactions were performed. For the FlowSOM analysis of activation markers expressed in memory T cells throughout time (6, 24, and 48h) in PA subjects, unsupervised clustering was performed based on the expression of CD25, CD69, OX40, CD137, and CD154 within memory CD4^+^ T cells. To characterize each cluster, mean fluorescence intensity (MFI) values were exported and corresponding pie charts were generated. All FlowSOM analyses were performed by using Cytobank platform and each set of samples were performed in triplicate to determine the consistency of the data obtained.

### 9. Statistical analysis

Data are reported as one individual and/or the mean ±/+ SEM, as specified. Statistical analyses were performed in Graphpad Prism v9 (GraphPad Software Inc., San Diego, CA) and TCR diversity in R using Immunarch ^55^. Significant differences between paired values of two groups were determined by Wilcoxon’s and Student’s t tests. Statistical significance between more than two groups accounting one or more variables was determined using mixed-effect analyses with Geisser-Greenhouse correction followed by Tukey’s and Dunnett’s multiple comparisons tests in GraphPad, as specified. Non-parametric data were analyzed using Kruskal-Wallis and Friedman tests followed by Dunn’s multiple comparisons test, as indicated. When noted, P values were corrected for false discovery rate (FDR). Differential cytokine abundance via LegendPlex assays were corrected for multiple comparisons using the Benjamini-Hochberg correction implemented in R via the p.adjust function. Post-hoc tests were only performed when main effects were significant. Differences were considered statistically significant if *P<0.05, **P<0.01, ***P<0.001, and ****P<0.0001. Unless otherwise specified, only significant P values are displayed.

#### 9.1. Statistics for diversity measures of TCR analysis

One-way ANOVAs were performed on all diversity measures using the aov function in R; post-hoc tests were performed using the TukeyHSD function. In all cases, the null hypothesis was that alpha diversity was not different across cellular subpopulation (sample_type: CD154^+^CD69^+^, CD137^+^CD154^-^, CD25^+^OX40^+^, and total memory CD45RA^-^ cells), after accounting for count depth. Formulas for the anova statistics were as follows:

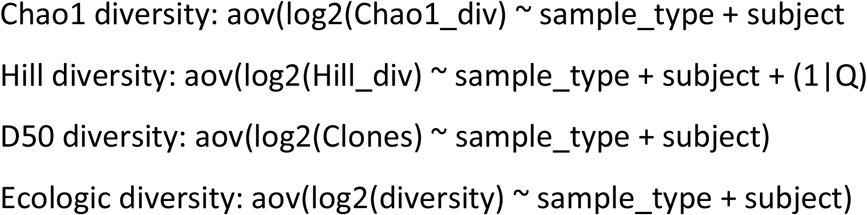

#### 9.2. Linear model selection for TCR analysis

The model residuals were all analyzed for deviation from the assumed normally distributed by Shapiro-Wilkes test of normality (shapiro.test function in R), and none were significantly different, therefore ensuring the fit of linear model assumptions. There is expected to be an effect of within-subject biology as well as a small amount of within-subject technical effects (how long blood sample sat in tube before cell isolation, etc). Additionally, the above described downsampling procedure produces a technical effect that is constant within a subject, but differs between subjects. Therefore, using subject as a covariate account for this subject-confounded technical effect alongside with any other subject specific effects. This also allows for a greater level of sensitivity because all subjects are not downsampled to the lowest level of any subject:sample_type pair. Each subject however is downsampled to the lowest level within their own subject_type measures (CD154^+^CD69^+^, CD137^+^CD154^-^, CD25^+^OX40^+^, and total memory CD45RA^-^ cells). Because there is only one measure per subject:sample_type pair, a fixed effect model is used for the subject factor rather than a repeated-measure random effect, as doing so collapses the model to invariance. The exception to this is the variable Q for Hill diversity. Hill diversity gives several values that show a curve for each subject:sample_type pair with Q on the X-axis; these “multiple-observations” must be taken into account as with a random effect as with a ‘repeated measure’ mixed-model style analysis.

## Data Availability

All data produced in the present study are available upon reasonable request to the authors

https://data.mendeley.com/datasets/tdxp6b8b64/draft?a=d027d3ba-67a9-40e0-8a32-6083f611b9e0

## Acknowledgements

We thank the Flow Cytometry Core and the Human Immune Monitoring Core at Mount Sinai. We thank C. Agashe and M. Mishoe for their expert technical assistance, and Jiaming Lin and Matthew Phelan for participant recruitment. We also thank the physicians, staff, allergic subjects, and their families of the Jaffe Food Allergy Institute. For the Consortium for Food Allergy Research 7 (CoFAR7), the following persons provided physician oversight, study coordination, and support: C. Bronchick, C. Chu, J. Fishman, D. Fitzgerald, D. Fleischer, J. French, E. Gibson, M. Groetch, D. Hamilton, P. Hauk, L. Herlihy, S. House, E. Kim, S. Leung, A. Liu, K. Mudd, T. Perry, R. Pesek, D. Robertson, J. Ross, A. Scurlock, J. Slinkard, P. Steele, L. Talarico, M. Taylor, and B. Vickery. We thank W. Davidson and M. Plaut for helpful contributions in mechanistic study planning. We thank A. Grishin and M. Masilamani for their leadership of laboratory studies of CoFAR7. We thank the staff of the clinical research units at each participating center, the Statistical and Clinical Coordinating Center, and K. Peyton, the SACCC Project Manager. We thank J. Poyser, Project Manager for CoFAR7 Program (National Institutes of Health [NIH]/National Institute of Allergy and Infectious Diseases [NIAID]). Finally, we thank the families who kindly participated.

## Author contributions

DLO and MCB designed this research. CA and MCL contributed to design B cell assays. SHS, SMJ, RAW, HAS, JW, AWB, and DYM oversaw clinical trial procedures (CoFAR7/CAFETERIA). DLO and CA performed the experiments. DLO, MCB, and ST analyzed data. DLO and MCB wrote the manuscript. All authors read and commented on the manuscript.

## Data and materials availability

All relevant are provided as source data in: https://data.mendeley.com/datasets/tdxp6b8b64/draft?a=d027d3ba-67a9-40e0-8a32-6083f611b9e0

TCRβ data and analyses are available in: https://bitbucket.org/scottyler892/tcr_analysis/src/master/

CoFAR7 data is available through ImmPORT (SDY1550). Any additional information required to reanalyze the data reported in this paper is available from the lead contact upon request.

Custom code was used for clonotype analysis on TCR sequencing data. Data and code are publicly available in the repository located at: https://bitbucket.org/scottyler892/tcr_analysis/src/master/.

