## SupplementalMaterial for "Allergen recognition by specific effector Th2 cells enables IL-2-dependent activation of regulatory T cell responses in humans"

**Supplementary Materials:**

***Fig. S1. Gating strategy for the identification of activated CD4$^+$ T cells and activated T cell populations in egg allergic subjects and non-allergic individuals.*** **A**. Gating strategy used for the identification of the activated populations (CD154$^+$CD69$^+$, CD137$^+$CD154$^-$, and CD25$^+$OX40$^+$) in the memory (CD45RA$^-$) CD4$^+$ T cells of PBMCs from donors. **B.** Percentage of the activated populations (CD154$^+$CD69$^+$, top left; CD137$^+$CD154$^-$, top right; CD25$^+$O40$^+$, bottom left) identified in memory CD4$^+$ T cells of PBMCs from non-allergic individuals unstimulated (CTRL), stimulated with crude peanut extract (CPE), and incubated with anti-human CD3/CD28 beads for 6h, 24h, and 48h (n=3). **C.** Percentage of the activated populations identified in memory CD4$^+$ T cells of PBMCs from egg allergic subjects unstimulated (CTRL) and stimulated with CPE or egg white proteins (EW) for 24h (n=5). In (B) and (C) each data point is one individual (mean ± SEM). In (B) statistical analysis by Friedman test followed by Dunn's multiple comparisons test and mixed-effect analysis followed by Tukey's multiple comparisons test in (C). *P<0.05, and **P<0.01.

***Fig. S2. Identification of regulatory T cells (Tregs) and gating strategy for fluorescence-activated cell-sorted (FACS) T cells.*** **A.** Representative flow cytometry plots of the Tregs (CD127$^-$Foxp3$^+$) within the three activated memory (CD45RA$^-$) CD4$^+$ T cell populations after stimulation of PBMCs from a peanut allergic subject with crude peanut extract for 6h, 24h, and 48h. **B.** Gating strategy used for the FACS-separation of the three activated populations (CD154$^+$CD69$^+$, CD137$^+$CD154$^-$, and CD25$^+$OX40$^+$) in the memory CD4$^+$ T cells magnetically pre-enriched from total PBMCs after stimulation. **C.** Representative flow cytometry plots of the Tregs (CD127$^-$Foxp3$^+$) within the memory CD4$^+$ T cell activated population (CD137$^+$CD154$^-$ and CD25$^+$OX40$^+$) after stimulation of PBMCs from a non-allergic donor with anti-human CD3/CD28 beads for 24h.

***Fig. S3. TCR diversity determined across peanut-activated CD4$^+$ T cell populations.*** **A**. $Log_2$ of the D50 estimate of diversity - this measures the number of clones that must be excluded before 50% of the dataset is removed for the given population. Lower D50 indicates greater clonotype homogeneity and lower diversity. Displayed are averages of 100 iterations, down-sampling to equivalence across classes within subjects. Individual points are colored by peanut allergic subjects. **B.** Hill numbers show that the lowest diversity belonging to CD154$^+$CD69$^+$ T cells induced after 6h of stimulation with crude peanut extract. **C.** Estimates of the percentage overlap of the clonotypes identified in the population of interest (x-axis) that appear in the noted box-plots, colorized by population type. Note that the population of interest is not shown because it will always equal 100%. Estimates shown are the results from 100 iterations of down-sampled versions of the dataset to fully account for differential sequencing depth.

***Fig. S4. Effect of peanut-induced cytokines in the generation of activated $CD4^+$ T cell populations and the role of IL-2 in the activation of regulatory T cells (Tregs) in peanut allergic (PA) subjects.*** **A.** Cytokine Secretion Assay (CSA) of activated populations after stimulation of PBMCs from peanut allergic (PA) subjects with crude peanut extract (CPE) for 2h-48h (n=4-12). **B.** T cell activation induced by stimulation of PBMCs from non-allergic individuals with a Th2 cytokine pool (IL-4+IL-5+IL-13) for 6h, 24h, and 48h. **C.** Neutralization of the induction of activated T cell populations in PBMCs from PA subjects (n=5) stimulated with CPE for 24h in presence of three different pools of anti-human cytokine blocking antibodies (legend at right). **D.** Neutralization of the induction of activated T cell populations in PBMCs from PA subjects (n=4-8) stimulated with CPE for 24h in presence of anti-human cytokine blocking antibodies and their isotypes controls. **E.** Percentage of activated populations ($CD154^+CD69^+$, $CD137^+CD154^-$, and $CD25^+OX40^+$) in non-CellTrace-labeled Tregs alone and pre-incubated with IL-2 (24h) after CPE-stimulation for 6h (n=5). **F.** IL-2 released by PBMCs cultured with purified Tregs alone or pre-incubated with IL-2 (24h) after CPE-stimulation for 6h (n=5). In (A) each point represents the mean ± SEM, one individual (mean ± SEM) in (B), (C), and (D), and one single individual in (E) and (F). In (A-D) statistical analysis by mixed-effect analysis with Geisser-Greenhouse correction followed by Tukey's multiple comparisons test. In (A), * expresses differences between $CD154^+CD69^+$ and $CD137^+CD154^-$ cells, # expresses differences between $CD154^+CD69^+$ and $CD25^+OX40^+$, and • expresses differences between $CD137^+CD154^-$ and $CD25^+OX40^+$ cells. In (E-F) statistical analysis by Wilcoxon's test. *,#,•P<0.05; **,##,••P<0.01; and ###,***P<0.001.

***Fig. S5.*** **In vivo** ***activation of regulatory T cells (Tregs) and Th2A cells before and after a double-blind placebo-controlled peanut challenge (DBPCPC) of peanut allergic (PA) subjects.*** **A.** Percentage of total $CD127_{low}CD25^+$ Tregs identified in PBMCs from PA subjects before (T0) and after (24h) a DBPCPC (n=7). **B.** Percentage of $CD69^+$ Th2A cells in PBMCs from PA subjects before (T0) and after (10d) a DBPCPC (n=9). **C.** Percentage of total $CD127_{low}CD25^+$ Tregs identified in

PBMCs from PA subjects before (T0) and after (10d) a DBPCPC (n=9). **D.** Percentage of CD25 expression in memory $CD127^{-}Foxp3^{+}$ T cells before (T0) and after (10d) a DBPCPC (n=9). Each data point is one individual. Statistical analysis by paired Student’s t test.

**Table S1.** Summarized clinical information of the pediatric subjects enrolled for this study.

| | **PA[1] subjects** | **EA[2] subjects** | **NA[3] donors** | **DBPCPC[4]** |
|---|---|---|---|---|
| **No.** | 58 | 5 | 3 | 21 |
| **Sex**, no. (M/F)[5] | 40/18 | 3/2 | 1/2 | 14/7 |
| **Age**, median (range) | 11 (4-18) | 18 (6-22) | 9 (6-14) | 7 (4-14) |
| **Race**, no. (H/AA/W/A/U)[6] | 4/3/28/6/17 | 0/1/2/0/2 | 0/0/2/0/1 | 0/1/14/6/0 |
| **Asthma**, no. (%) | 39 (73.6) | 5 (100) | N/A[7] | 1 (4.8) |
| **Allergic rhinitis**, no. (%) | 39 (73.6) | 5 (100) | N/A | 12 (57.1) |
| **Atopic dermatitis**, no. (%) | 43 (81.2) | 2 (40) | N/A | 12 (57.1) |
| **Tree nut allergy**, no. (%) | 50 (94.3) | 4 (80) | N/A | 10 (47.6) |
| **Other food allergy**, no. (%) | 32 (60.4) | 5 (100) | N/A | 11 (52.4) |
| **Total IgE** median (range, kUA/L) | 7 21.0 (64.1-9664.2) | 808.0 (451.0-1952.0) | 10.0 (1.50-30.0) | 222.0 (32.0-477.0) |
| **Peanut sIgE**[8] median (range, kUA/L) | 91.1 (0.4-806.1) | ND[9] | ND | 7.2 (0.4-36.6) |
| **Arah1 sIgE** median (range, kUA/L) | 20.4 (0.1-100.0) | N/A | N/A | N/A |
| **Arah2 sIgE** median (range, kUA/L) | 56.2 (0.3-100.0) | N/A | N/A | 4.1 (0.1-20.9) |
| **Egg sIgE** median (range, kUA/L) | N/A | 17.7 (0.9-100.0) | ND | N/A |

[1]PA: Peanut allergic. [2]EA: Egg allergic. [3]NA: Non-allergic. [4]DBPCPC: PA subjects undergoing a double-blind placebo-controlled peanut challenge. [5]M: Male, F: Female. [6]H: Hispanic/Latino, AA: African American, W: White, A: Asian, U: unknown or with more than one race. [7]N/A: not available. [8]sIgE: Specific-IgE. [9]ND: non-detected.

**Table S2.** List of anti-human antibodies and viability dye used for the identification of activated populations as well as their phenotypical and functional characterization by conventional flow cytometry using a LSR Fortessa device (BD Bioscience).

| **Target** | **Conjugate** | **Clone** | **Company** | **Reference** | **Dilution** |
|---|---|---|---|---|---|
| Foxp3 | AF647 | 206D | BioLegend | 320114 | 1:50 |
| CCR7 | AF700 | G043H7 | BioLegend | 353244 | 1:40 |
| CD3 | APC-efluor780 | SK7 | Invitrogen | 47-0036-42 | 1:100 |
| CD69 | Pacific blue | FN50 | BioLegend | 310920 | 1:77 |
| HLA-DR | BV510 | L243 | BioLegend | 307646 | 1:25 |
| CD25 | BV605 | BC96 | BioLegend | 302632 | 1:33 |
| OX40 | BV711 | Ber-ATC35 | BioLegend | 350030 | 1:33 |
| CD45RA | BV785 | HI100 | BioLegend | 304140 | 1:66 |
| CD154 | PE | 24-31 | Invitrogen | 12-1548-42 | 1:25 |
| CD137 | PE-Cy5 | 4B4-1 | BioLegend | 309808 | 1:33 |
| CD27 | PerCP-Cy5.5 | M-T271 | BioLegend | 356408 | 1:33 |
| Ki67 | AF488 | Ki-67 | BioLegend | 350508 | 1:33 |
| CD4 | PE-Cy7 | RPA-T4 | Invitrogen | 25-0049-42 | 1:66 |
| CD27 | FITC | M-T271 | BioLegend | 356403 | 1:66 |
| CD4 | PE-CF594 | RPA-T4 | BD Horizon | 562316 | 1:80 |
| CD27 | PerCP-efluor710 | O323 | Invitrogen | 46-0279-42 | 1:50 |
| CCR4 | PerCP-Cy5.5 | 161 | BD Pharmingen | 560726 | 1:25 |
| CD161 | APC | HP-3610 | Invitrogen | 17-1619-42 | 1:33 |
| CCR7 | BV785 | G043H7 | BioLegend | 353230 | 1:33 |
| CD25 | AF700 | BC96 | BioLegend | 302622 | 1:33 |
| CCR7 | BV510 | G043H7 | BioLegend | 353232 | 1:33 |
| CD154 | FITC | 24-31 | Invitrogen | 11-1548-42 | 1:25 |
| Live/Dead | For UV excitation | N/A[1] | Themo Fisher | L34962 | 1:1000 |

[1]N/A: not applicable.

**Table S3.** List of anti-human antibodies and viability dye used for the phenotypical and functional characterization of the peanut-induced CD4+ T cells by spectral flow cytometry using a 4-laser Cytek™ Aurora device (Cytek Biosciences).

| Target | Conjugate | Clone | Company | Reference | Dilution |
|---|---|---|---|---|---|
| TIGIT | BV421 | A151536 | BioLegend | 372710 | 1:50 |
| CD62L | SB 436 | DREG-56 | Invitrogen | 62-0629-42 | 1:50 |
| IRF4 | eFluor450 | 3E4 | Invitrogen | 48-9858-82 | 1:120 |
| CD127 | BV480 | HIL-7R-H21 | BD Horizon | 566101 | 1:50 |
| TCR-$\beta$ | eFluor506 | H57-597 | Invitrogen | 69-5961-82 | 1:50 |
| PD1 | BV510 | EH12.2H7 | BioLegend | 329932 | 1:75 |
| CD45RA | BV570 | HI100 | BioLegend | 304132 | 1:100 |
| CCR6 | BV605 | G034E3 | BioLegend | 353420 | 1:75 |
| CD25 | BV650 | BC96 | BioLegend | 302634 | 1:75 |
| CD27 | Qdot655 | CLB-2711 | Invitrogen | Q10066 | 1:300 |
| OX40 | BV711 | Ber-ACT35 | BioLegend | 350030 | 1:50 |
| CD3 | BV750 | SK7 | BioLegend | 344846 | 1:120 |
| CCR7 | BV785 | G043H7 | BioLegend | 353230 | 1:75 |
| CXCR5 | AF488 | RF8B2 | BD Pharmingen | 558112 | 1:50 |
| CD4 | AF532 | RPA-T4 | Invitrogen | 58-0049-42 | 1:100 |
| HLA-DR | PerCP | L243 | BioLegend | 307628 | 1:60 |
| ICOS | PerCP-Cy5.5 | C398.4A | BioLegend | 313518 | 1:60 |
| CD69 | PerCP-efluor710 | FN50 | Invitrogen | 46-0699-42 | 1:80 |
| CD154 | PE | 24-31 | Invitrogen | 12-1548-42 | 1:40 |
| CXCR3 | Pe-efluor610 | CEW33D | Invitrogen | 61-1839-42 | 1:75 |
| CD137 | PE-Cy5 | 4B4-1 | BioLegend | 309808 | 1:75 |
| Foxp3 | PE-Cy7 | PCH101 | Invitrogen | 25-4776-42 | 1:80 |
| BCL6 | APC | 7D1 | BioLegend | 358506 | 1:60 |
| CCR4 | AF647 | L291H4 | BioLegend | 359404 | 1:75 |
| CD49d[1] | Purified | 9F10 | Invitrogen | 14-0499-82 | 1:75 |
| CD161 | AF700 | HP-3610 | BioLegend | 339942 | 1:80 |
| CRTH2 | APC-Vio770 | REA598 | Miltenyi Biotec | 130-119-623 | 1:100 |
| Live/Dead | Zombie NIR | N/A[2] | BioLegend | 423106 | 1:10000 |

[1]Labeled with APC-Cy5.5 using a commercial conjugation kit from abcam (reference: ab102855).[2]N/A: not applicable.

**Table S4.** List of anti-human antibodies and viability dye used for the evaluation of the *in vivo* activation of memory CD4+ T cells in peanut allergic subjects undergoing a double-blind placebo-controlled peanut challenge by spectral flow cytometry using a 5-laser Cytek™ Aurora device (Cytek Biosciences).

| Target | Conjugate | Clone | Company | Reference | Dilution |
|---|---|---|---|---|---|
| CXCR3 | BV421 | G025H7 | BioLegend | 353716 | 1:75 |
| CD3 | SB436 | SK7 | Invitrogen | 62-0036-42 | 1:100 |
| IRF4 | eFluor450 | 3E4 | Invitrogen | 48-9858-82 | 1:100 |
| ICOS | BV480 | 7E.17G9 | BD Biosciences | 746248 | 1:75 |
| CCR4 | BV510 | L29IH4 | BioLegend | 359416 | 1:60 |
| CD45RA | BV570 | HI100 | BioLegend | 304132 | 1:100 |
| CCR6 | BV605 | G034E3 | BioLegend | 353420 | 1:75 |
| CD25 | BV650 | BC96 | BioLegend | 302634 | 1:75 |
| OX40 | BV711 | Ber-ACT35 | BioLegend | 350030 | 1:50 |
| PD1 | BV750 | EH12.2H7 | BioLegend | 329966 | 1:75 |
| CD27 | BV785 | O323 | BioLegend | 302832 | 1:75 |
| CD154 | FITC | 24-31 | Invitrogen | 11-1548-42 | 1:50 |
| CD4 | AF532 | RPA-T4 | Invitrogen | 58-0049-42 | 1:100 |
| HLA-DR | PerCP | L243 | BioLegend | 307628 | 1:60 |
| LAP | PerCP-Cy5.5 | S20006A | BioLegend | 349612 | 1:75 |
| CD69 | PerCP-efluor710 | FN50 | Invitrogen | 46-0699-42 | 1:80 |
| Ki67 | PE | SolA15 | Invitrogen | 12-5698-82 | 1:75 |
| CD49d | PE-Dazzle594 | 9F10 | BioLegend | 304326 | 1:100 |
| CD137 | PE-Cy5 | 4B4-1 | BioLegend | 309808 | 1:75 |
| Foxp3 | PE-Cy7 | PCH101 | Invitrogen | 25-4776-42 | 1:80 |
| CD127 | APC | A019D5 | BioLegend | 351316 | 1:80 |
| CXCR5 | AF647 | RF8B2 | BD Biosciences | 558113 | 1:75 |
| CD161 | AF700 | HP-3610 | BioLegend | 339942 | 1:80 |
| CRTH2 | APC-Vio770 | REA598 | Miltenyi Biotec | 130-119-623 | 1:100 |
| CD71 | BUV563 | M-A712 | BD Biosciences | 749296 | 1:100 |
| CD38 | BUV615 | HIT2 | BD Biosciences | 751138 | 1:100 |
| CCR7 | BUV737 | 3D12 | BD Biosciences | 741786 | 1:100 |
| Live/Dead | For UV excitation | N/A[1] | Themo Fisher | L34962 | 1:5000 |

[1]N/A: not applicable.

**Table S5.** List of anti-human antibodies and viability dye used for fluorescence-activated cell-sorted (FACS) T cell populations after activation by peanut allergens by using a FACS Aria II device (BD Bioscience).

| Target | Conjugate | Clone | Company | Reference | Dilution |
|---|---|---|---|---|---|
| CD3 | APC-efluor780 | SK7 | Invitrogen | 47-0036-42 | 1:75 |
| OX40 | APC-Cy7 | Ber-ACT35 | BioLegend | 350022 | 1:50 |
| CD154 | FITC | 24-31 | Invitrogen | 11-1548-42 | 1:25 |
| CD69 | Pacific blue | FN50 | BioLegend | 310920 | 1:80 |
| CD25 | BV785 | BC96 | BioLegend | 302638 | 1:75 |
| CD137 | PE | 4B4-1 | BioLegend | 309804 | 1:30 |
| CD4 | PE-CF594 | RPA-T4 | BD Horizon | 562316 | 1:50 |
| CD45RA | PE-Cy7 | HI100 | Invitrogen | 25-0458-42 | 1:80 |
| Live/Dead | For UV excitation | N/A[1] | Themo Fisher | L34962 | 1:5000 |

[1]N/A: not applicable.

**Table S6.** List of anti-human antibodies and viability dye used for the analyses of the immunosuppression assays by conventional flow cytometry using a using a CytoFLEX device (Beckman Coulter).

| Target | Conjugate | Clone | Company | Reference | Dilution |
|---|---|---|---|---|---|
| CD4 | BV605 | OKT4 | BioLegend | 317438 | 1:100 |
| CD3 | PE-CF594 | UCHT1 | BD Horizon | 562310 | 1:100 |
| CD127 | PE-Cy7 | A019D5 | BioLegend | 351320 | 1:80 |
| Foxp3 | APC | PCH101 | Invitrogen | 17-4776-42 | 1:80 |
| CD25 | AF700 | BC96 | BioLegend | 302622 | 1:100 |
| CD45RA | PerCP | HI100 | BioLegend | 204156 | 1:100 |
| Live/Dead | For 405 nm excitation | N/A[1] | Themo Fisher | L34962 | 1:1000 |

[1]N/A: not applicable.

**Table S7.** List of anti-human antibodies, viability dye and commercial detection kits used in the cytokine secretion assay experiments for the identification of cytokine-secreting activated CD4+ T cells by conventional flow cytometry using a LSR Fortessa device (BD Bioscience).

| **Target** | **Conjugate** | **Clone** | **Company** | **Reference** | **Dilution** |
|---|---|---|---|---|---|
| CD154 | FITC | 24-31 | Invitrogen | 11-1548-42 | 1:25 |
| CD25 | AF700 | BC96 | BioLengend | 302622 | 1:40 |
| CD3 | APC-efluor780 | SK7 | Invitrogen | 47-0036-42 | 1:66 |
| CD27 | PerCP-efluor710 | O323 | Invitrogen | 46-0279-42 | 1:50 |
| CCR7 | BV510 | G043H7 | BioLegend | 353232 | 1:33 |
| CD69 | Pacific Blue | FN50 | BioLegend | 310920 | 1:100 |
| OX40 | BV711 | Ber-ATC35 | BioLegend | 350030 | 1:33 |
| CD45RA | BV785 | HI100 | BioLegend | 304140 | 1:66 |
| CD4 | PE-CF594 | RPA-T4 | BD Horizon | 562316 | 1:50 |
| CD137 | PE-Cy5 | 4B4-1 | BioLegend | 309808 | 1:33 |
| CD127 | PE-Cy7 | eBioRDR5 | Invitrogen | 25-1278-42 | 1:25 |
| IL-13 | PE | N/A[1] | Miltenyi | 130-093-479 | REC[2] |
| IL-10 | APC | N/A | Miltenyi | 130-091-761 | REC |
| IL-4 | PE | N/A | Miltenyi | 130-054-102 | REC |
| IL-5 | APC | N/A | Miltenyi | 130-091-624 | REC |
| IL-2 | PE | N/A | Miltenyi | 130-090-487 | REC |
| IFN-$\gamma$ | APC | N/A | Miltenyi | 130-090-762 | REC |
| TNF-$\alpha$ | APC | N/A | Miltenyi | 130-091-267 | REC |
| Live/Dead | For UV excitation | N/A | Themo Fisher | L34962 | 1:1000 |

[1]N/A: not applicable. [2]REC: concentration recommended by the manufacturer.

**Table S8.** List of anti-human antibodies and viability dye used in the intracellular cytokine secretion (ICS) experiments for the identification of cytokine-producing activated CD4+ T cells by spectral flow cytometry using a 4-laser Cytek™ Aurora device (Cytek Biosciences).

| Target | Conjugate | Clone | Company | Reference | Dilution |
|---|---|---|---|---|---|
| IL-5 | BV421 | TRFK5 | BioLegend | 504311 | 1:45 |
| IL-25 | AF405 | 182203 | R&D Systems | IC1258V | 1:40 |
| IL-13 | Horizon V450 | JE510-SA2 | BD Horizon | 561158 | 1:50 |
| CD127 | BV480 | HIL-7R-H21 | BD Horizon | 566101 | 1:50 |
| IL-17A | eFluor506 | eBio64DEC17 | Invitrogen | 69-7179-42 | 1:40 |
| CCR4 | BV510 | L29IH4 | BioLegend | 359416 | 1:60 |
| CD45RA | BV570 | HI100 | BioLegend | 304132 | 1:100 |
| CCR6 | BV605 | G034E3 | BioLegend | 353420 | 1:75 |
| IL-2 | BV650 | MQ1-17H12 | BioLegend | 500334 | 1:60 |
| OX40 | BV711 | Ber-ACT35 | BioLegend | 350030 | 1:50 |
| CD3 | BV750 | SK7 | BioLegend | 344846 | 1:120 |
| CD25 | BV785 | BC96 | BioLegend | 302638 | 1:75 |
| CD154 | FITC | 24-31 | Invitrogen | 11-1548-42 | 1:40 |
| CD4 | AF532 | RPA-T4 | Invitrogen | 58-0049-42 | 1:100 |
| TNF-α | PerCP | Mab11 | BioLegend | 502924 | 1:75 |
| GM-CSF | PerCP-Cy5.5 | BVD2-21C11 | BioLegend | 502312 | 1:50 |
| CD69 | PerCP-efluor710 | FN50 | Invitrogen | 46-0699-42 | 1:80 |
| IL-21 | PE | 3A3-N2 | BioLegend | 513004 | 1:50 |
| IL-10 | PE-CF594 | JES3-19F1 | BD Horizon | 562400 | 1:60 |
| CD137 | PE-Cy5 | 4B4-1 | BioLegend | 309808 | 1:75 |
| IL-9 | PE-Cy7 | MH9A4 | BioLegend | 507612 | 1:50 |
| CCR3 | APC | 5E8 | BioLegend | 310708 | 1:75 |
| IL-4 | AF647 | MP4-25D2 | BioLegend | 500818 | 1:60 |
| CD38 | APC-Cy5.5 | HIT2 | Invitrogen | MHCD3819 | 1:30 |
| IFN-γ | AF700 | B27 | BD Pharmingen | 557995 | 1:75 |
| CRTH2 | APC-Vio770 | REA598 | Miltenyi Biotec | 130-119-623 | 1:100 |
| Live/Dead | Zombie NIR | N/A[1] | BioLegend | 423106 | 1:10000 |

[1] N/A: not applicable.

**Table S9.** List of anti-human antibodies and viability dye used for the identification of activated populations in the experiments determining the conditions of induction and neutralization of the bystander induced subsets analyzed by conventional flow cytometry using a LSR Fortessa device (BD Bioscience).

| **Target** | **Conjugate** | **Clone** | **Company** | **Reference** | **Dilution** |
|---|---|---|---|---|---|
| CD69 | Pacific blue | FN50 | BioLegend | 310920 | 1:80 |
| CD3 | APC-efluor780 | SK7 | Invitrogen | 47-0036-42 | 1:100 |
| CD4 | PE-CF594 | RPA-T4 | BD Horizon | 562316 | 1:80 |
| CD154 | PE | 24-31 | Invitrogen | 12-1548-42 | 1:40 |
| CD137 | PE-Cy7 | 4B4-1 | BioLegend | 309818 | 1:60 |
| Foxp3 | APC | PCH101 | Invitrogen | 17-4776-42 | 1:80 |
| CD25 | AF700 | BC96 | BioLegend | 302622 | 1:100 |
| OX40 | FITC | Ber-ACT35 | BioLegend | 350006 | 1:80 |
| CD45RA | PerCP | HI100 | BioLegend | 204156 | 1:100 |
| Live/Dead | For UV excitation | N/A[1] | Themo Fisher | L34962 | 1:1000 |

[1]N/A: not applicable.

**Table S10.** List of anti-human antibodies and viability dye used in the experiments performed for the determination of the role of IL-2 in the functional activation of Tregs by spectral flow cytometry using a 5-laser Cytek™ Aurora device (Cytek Biosciences).

| Target | Conjugate | Clone | Company | Reference | Dilution |
|---|---|---|---|---|---|
| CXCR3 | BV421 | G025H7 | BioLegend | 353716 | 1:75 |
| CD3 | SB436 | SK7 | Invitrogen | 62-0036-42 | 1:100 |
| CCR4 | BV510 | L29IH4 | BioLegend | 359416 | 1:60 |
| CD45RA | BV570 | HI100 | BioLegend | 304132 | 1:100 |
| CCR6 | BV605 | G034E3 | BioLegend | 353420 | 1:75 |
| CD25 | BV650 | BC96 | BioLegend | 302634 | 1:50 |
| OX40 | BV711 | Ber-ACT35 | BioLegend | 350030 | 1:50 |
| CCR7 | BV750 | G043H7 | BioLegend | 353254 | 1:100 |
| CD27 | BV785 | O323 | BioLegend | 302832 | 1:75 |
| CD154 | FITC | 24-31 | Invitrogen | 11-1548-42 | 1:50 |
| CD4 | AF532 | RPA-T4 | Invitrogen | 58-0049-42 | 1:100 |
| HLA-DR | PerCP | L243 | BioLegend | 307628 | 1:60 |
| CD69 | PerCP-efluor710 | FN50 | Invitrogen | 46-0699-42 | 1:80 |
| Ki67 | PE | SolA15 | Invitrogen | 12-5698-82 | 1:75 |
| CD137 | PE-Cy5 | 4B4-1 | BioLegend | 309808 | 1:75 |
| Foxp3 | PE-Cy7 | PCH101 | Invitrogen | 25-4776-42 | 1:80 |
| GATA3 | APC | 16E10A23 | BioLegend | 653806 | 1:75 |
| CD127 | AF700 | A019D5 | BioLegend | 351344 | 1:75 |
| CRTH2 | APC-Vio770 | REA598 | Miltenyi Biotec | 130-119-623 | 1:100 |
| Live/Dead | For UV excitation | N/A[1] | Themo Fisher | L34962 | 1:5000 |

[1]N/A: not applicable.

**Table S11.** List of anti-human antibodies and viability dye used for fluorescence-activated cell-sorted (FACS) depletion of Tregs by using a FACS Aria II device (BD Bioscience).

| Target | Conjugate | Clone | Company | Reference | Dilution |
|---|---|---|---|---|---|
| CD3 | APC-efluor780 | SK7 | Invitrogen | 47-0036-42 | 1:75 |
| CD4 | APC | RPA-T4 | Invitrogen | 17-0049-42 | 1:50 |
| CD25 | AF700 | BC96 | BioLegend | 302622 | 1:100 |
| CD127 | BV785 | A7R34 | BioLegend | 135037 | 1:100 |
| Live/Dead | For UV excitation | N/A[1] | Themo Fisher | L34962 | 1:5000 |

[1]N/A: not applicable.

### Figure S1

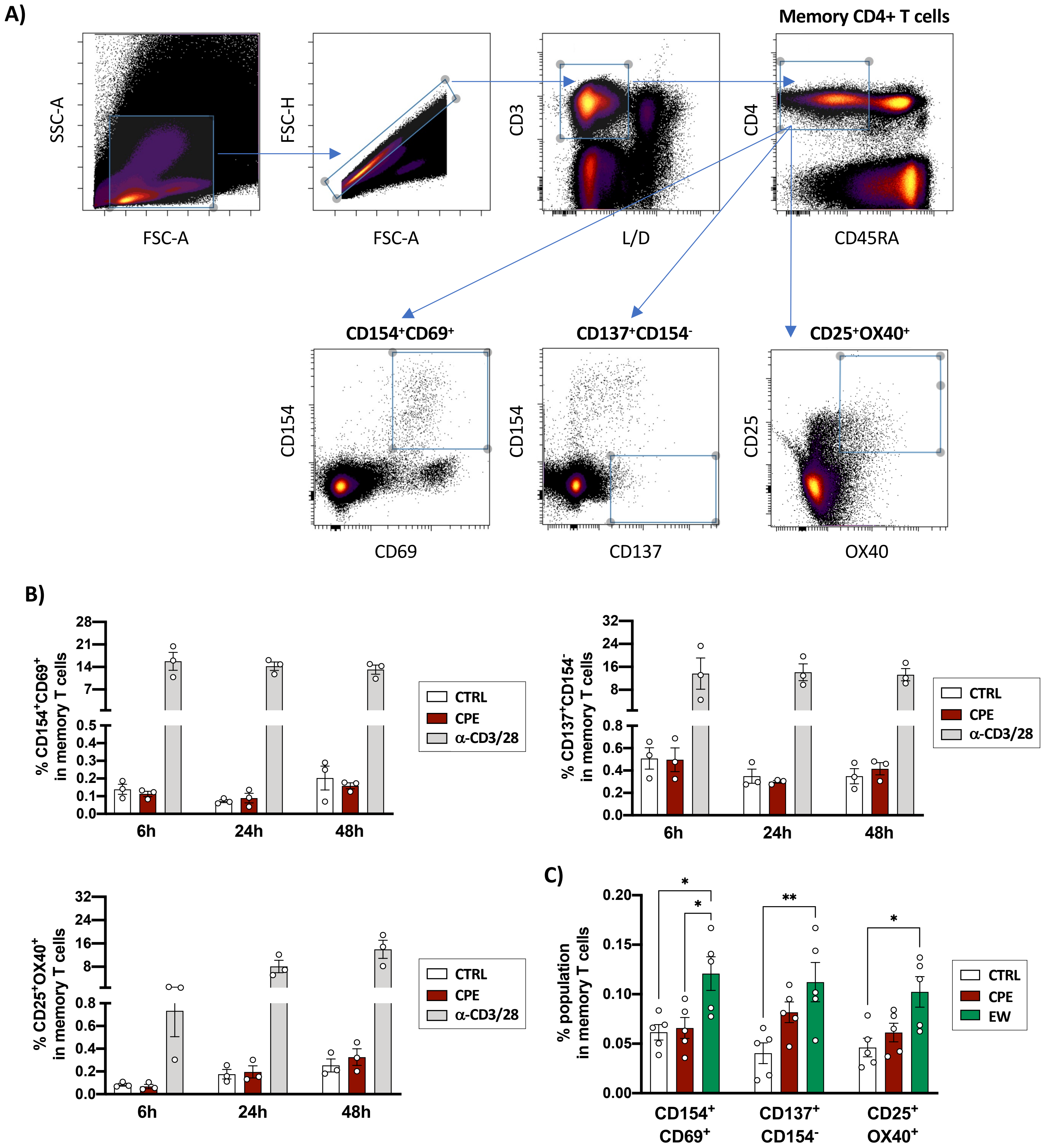

A)
Memory CD4+ T cells
SSC-A
FSC-A
FSC-H
FSC-A
CD3
L/D
CD4
CD45RA
CD154⁺CD69⁺
CD137⁺CD154⁻
CD25⁺OX40⁺
CD154
CD69
CD154
CD137
CD25
OX40
B)
% CD154⁺CD69⁺ in memory T cells
% CD137⁺CD154⁻ in memory T cells
% CD25⁺OX40⁺ in memory T cells
6h
24h
48h
CTRL
CPE
α-CD3/28
C)
% population in memory T cells
CD154⁺ CD69⁺
CD137⁺ CD154⁻
CD25⁺ OX40⁺
CTRL
CPE
EW

### Figure S2

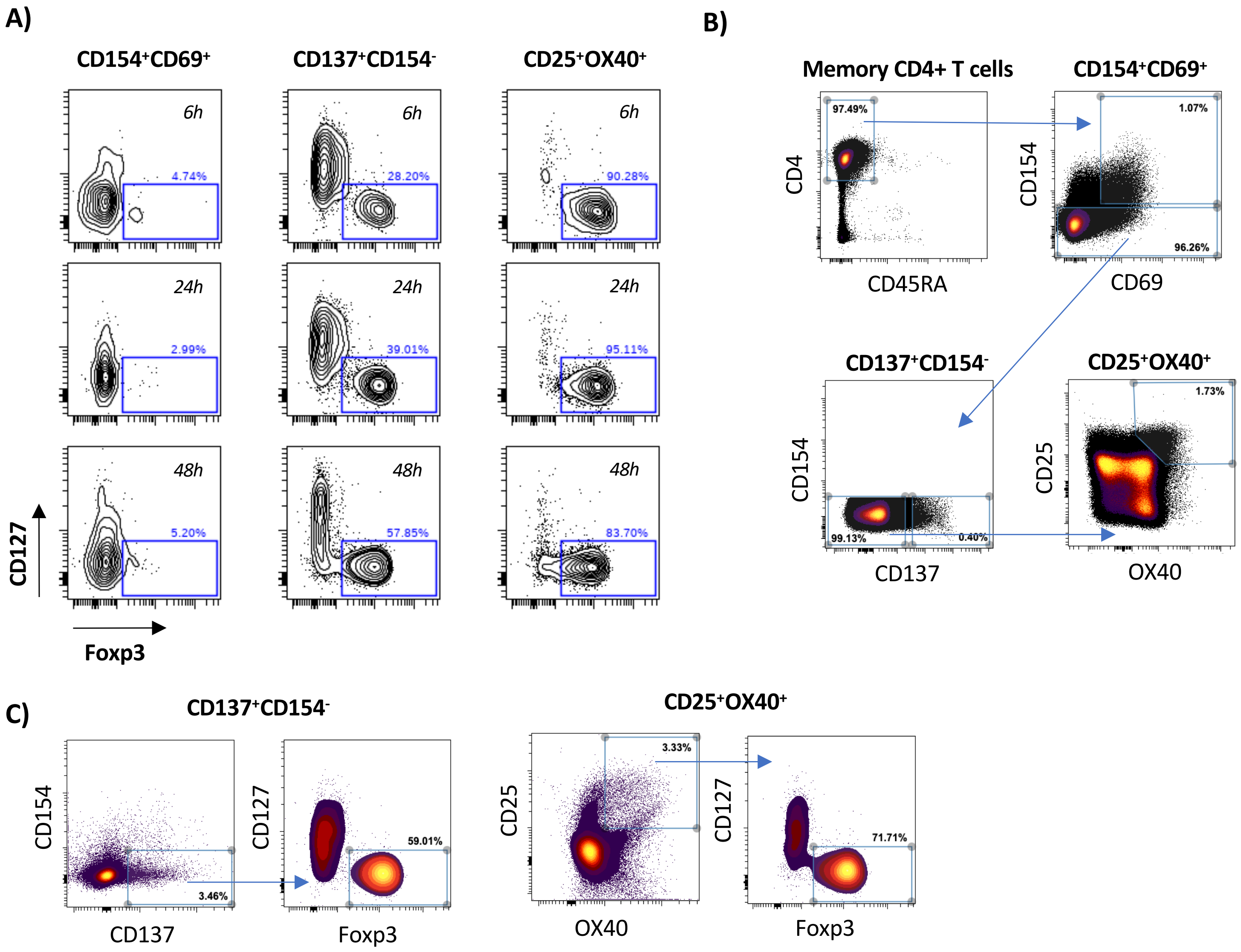
A)
CD154⁺CD69⁺
CD137⁺CD154⁻
CD25⁺OX40⁺
6h
24h
48h
4.74%
28.20%
90.28%
2.99%
39.01%
95.11%
5.20%
57.85%
83.70%
CD127
Foxp3
B)
Memory CD4+ T cells
CD154⁺CD69⁺
97.49%
CD4
CD45RA
1.07%
96.26%
CD154
CD69
CD137⁺CD154⁻
CD25⁺OX40⁺
99.13%
0.40%
CD154
CD137
1.73%
CD25
OX40
C)
CD137⁺CD154⁻
CD25⁺OX40⁺
CD154
3.46%
CD137
CD127
59.01%
Foxp3
CD25
3.33%
OX40
CD127
71.71%
Foxp3

#### Figure S3

A)

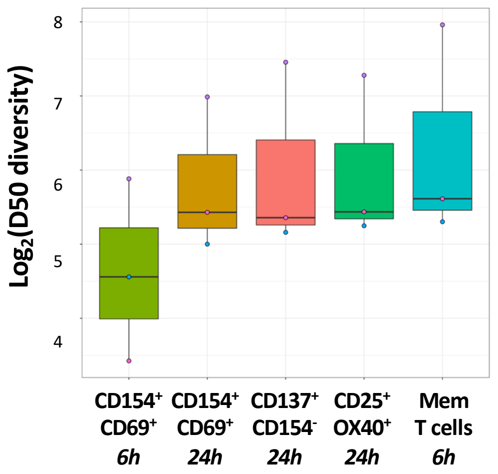


B)

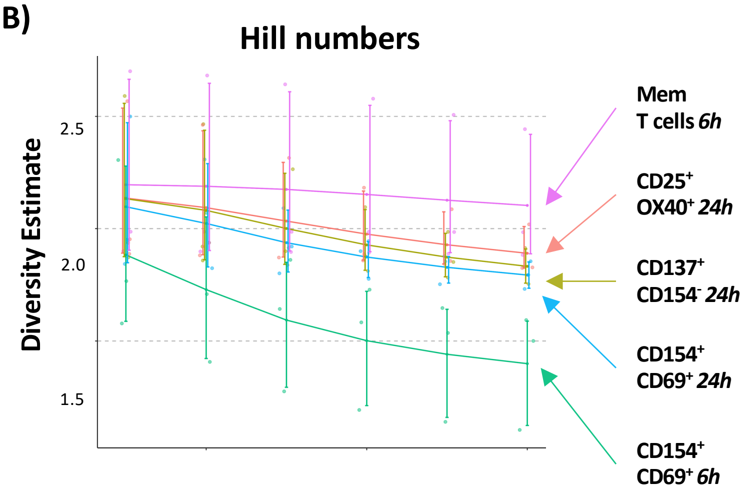


C)

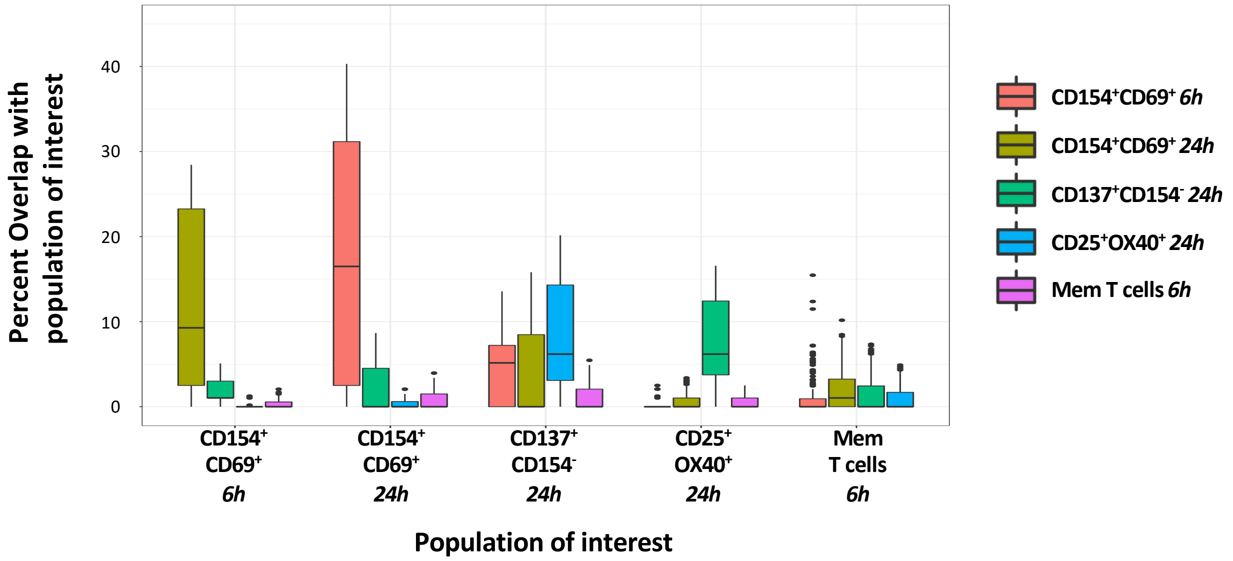

#### Figure S4

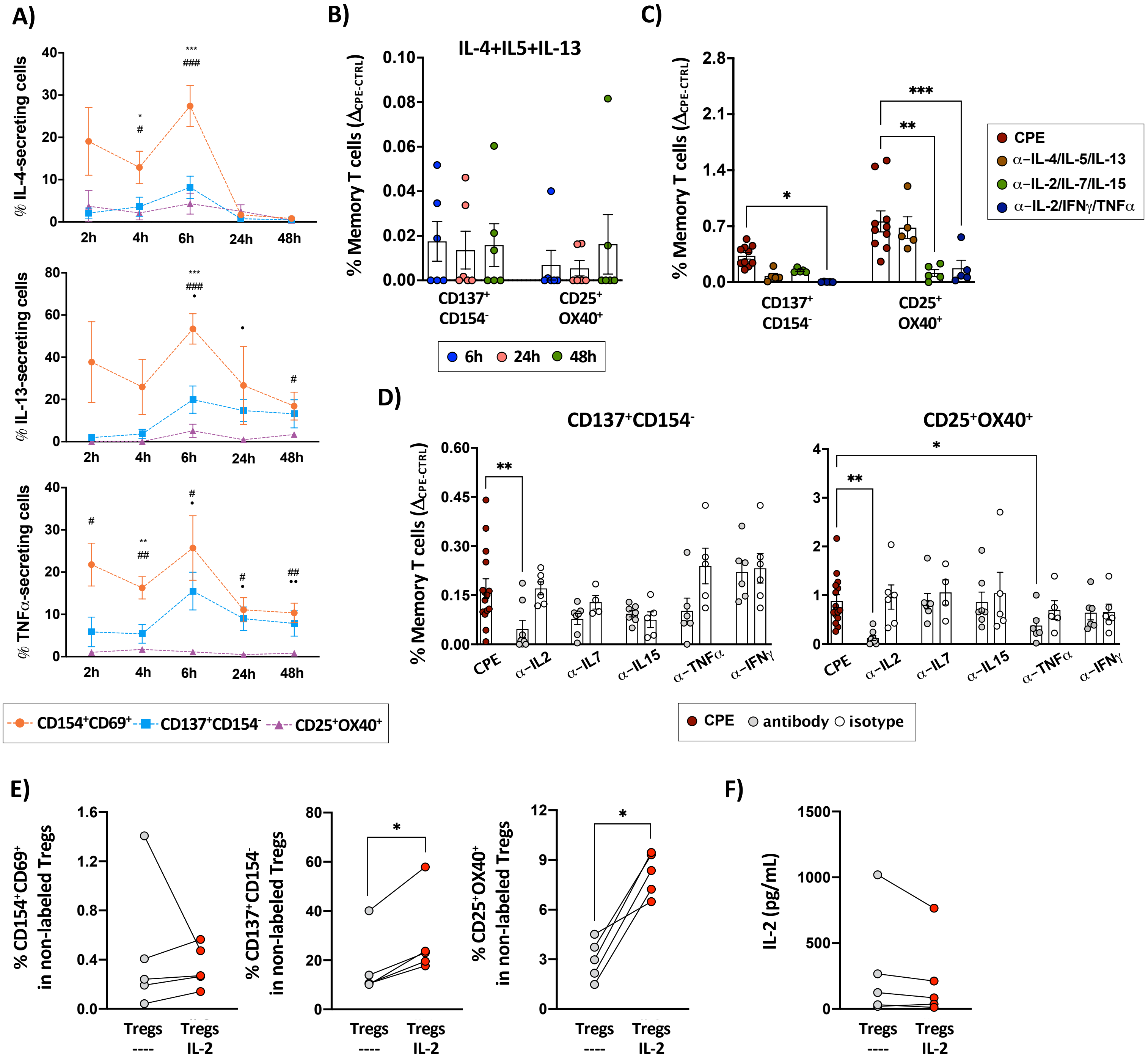

A)
% IL-4-secreting cells
% IL-13-secreting cells
% TNFα-secreting cells
2h 4h 6h 24h 48h
CD154+CD69+ CD137+CD154- CD25+OX40+
B)
IL-4+IL5+IL-13
% Memory T cells (ΔCPE-CTRL)
CD137+ CD154-
CD25+ OX40+
6h 24h 48h
C)
CPE
α-IL-4/IL-5/IL-13
α-IL-2/IL-7/IL-15
α-IL-2/IFNγ/TNFα
D)
CD137+CD154-
CD25+OX40+
CPE α-IL2 α-IL7 α-IL15 α-TNFα α-IFNγ
CPE antibody isotype
E)
% CD154+CD69+ in non-labeled Tregs
% CD137+CD154- in non-labeled Tregs
% CD25+OX40+ in non-labeled Tregs
Tregs ----
Tregs IL-2
F)
IL-2 (pg/mL)

### Figure S5

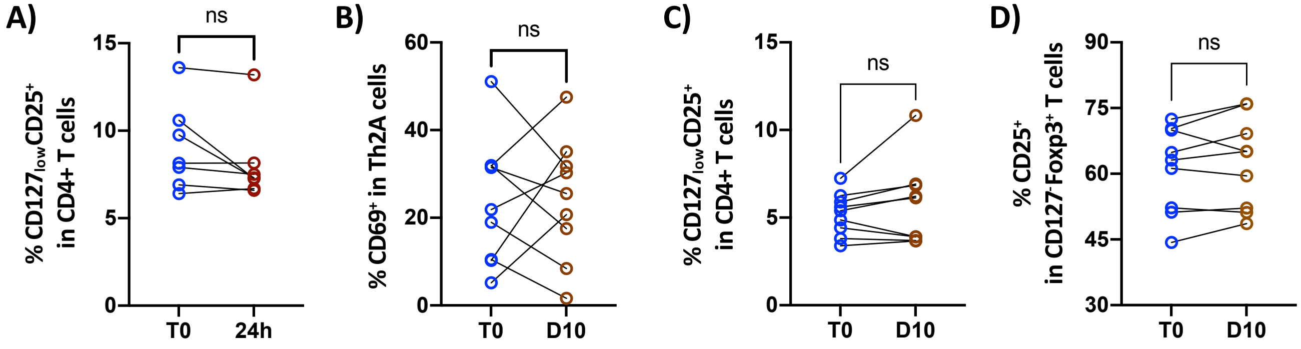

A)
ns
% CD127lowCD25+ in CD4+ T cells
15
10
5
0
T0
24h
B)
ns
% CD69+ in Th2A cells
60
40
20
0
T0
D10
C)
ns
% CD127lowCD25+ in CD4+ T cells
15
10
5
0
T0
D10
D)
ns
% CD25+ in CD127-Foxp3+ T cells
90
75
60
45
30
T0
D10
